## Supplementary material for "High prevalence of an alpha variant lineage with a premature stop codon in ORF7a in Iraq, winter 2020-2021": Table S1

**Supplementary materials**

**Table S1**: Acknowledgement to the contributors of the SARS-CoV-2 sequences used in this study.

We gratefully acknowledge the following Authors from the Originating laboratories responsible for obtaining the specimens and the Submitting laboratories where genetic sequence data were generated and shared via the GISAID Initiative, on which this research is based.

All submitters of data may be contacted directly via www.gisaid.org

| Virus name | Accession No. | Originating laboratory | Submitting laboratory | Authors |
|---|---|---|---|---|
| **hCoV-19/Wuhan/WIV04/2019** | EPI_ISL_402124 | Wuhan Jinyintan Hospital | Wuhan Institute of Virology, Chinese Academy of Sciences | Peng Zhou, Xing-Lou Yang, Ding-Yu Zhang, Lei Zhang, Yan Zhu, Hao-Rui Si, Zhengli Shi |
| **hCoV-19/Iraq/Samawa-1/2021** | EPI_ISL_1524330 | Biology Department, College of Science, Al Muthanna University and Public Health Laboratory, Al-Muthanna Health Directorate | Department of Virology, Faculty of Medicine, University of Helsinki, Helsinki, Finlan | Nihad Al-Rashedi, Hussein Alburkat, Murad Munahi, Alaa Hameed, Ali Jasim, Olli Vapalahti, Tarja Sironen,Teemu Smura |
| **hCoV-19/Iraq/Samawa-2/2021** | EPI_ISL_1524331 | Biology Department, College of Science, Al Muthanna University and Public Health Laboratory, Al-Muthanna Health Directorate | Department of Virology, Faculty of Medicine, University of Helsinki, Helsinki, Finlan | Nihad Al-Rashedi, Hussein Alburkat, Murad Munahi, Alaa Hameed, Ali Jasim, Olli Vapalahti, Tarja Sironen,Teemu Smura |
| **hCoV-19/Iraq/Samawa-3/2021** | EPI_ISL_1524332 | Biology Department, College of Science, Al Muthanna University and Public Health Laboratory, Al-Muthanna Health Directorate | Department of Virology, Faculty of Medicine, University of Helsinki, Helsinki, Finlan | Nihad Al-Rashedi, Hussein Alburkat, Murad Munahi, Alaa Hameed, Ali Jasim, Olli Vapalahti, Tarja Sironen,Teemu Smura |
| **hCoV-19/Iraq/Samawa-** | EPI_ISL_1524333 | Biology Department, College of Science, Al Muthanna University and Public Health | Department of Virology, Faculty of Medicine, University of Helsinki, | Nihad Al-Rashedi, Hussein Alburkat, Murad Munahi, Alaa Hameed, Ali Jasim, Olli |

| | | | | |
|---|---|---|---|---|
| **4/2021** | | Laboratory, Al-Muthanna Health Directorate | Helsinki, Finlan | Vapalahti, Tarja Sironen,Teemu Smura |
| **hCoV-19/Iraq/Samawa-5/2021** | EPI_ISL_1524334 | Biology Department, College of Science, Al Muthanna University and Public Health Laboratory, Al-Muthanna Health Directorate | Department of Virology, Faculty of Medicine, University of Helsinki, Helsinki, Finlan | Nihad Al-Rashedi, Hussein Alburkat, Murad Munahi, Alaa Hameed, Ali Jasim, Olli Vapalahti, Tarja Sironen,Teemu Smura |
| **hCoV-19/Iraq/Samawa-6/2021** | EPI_ISL_1524335 | Biology Department, College of Science, Al Muthanna University and Public Health Laboratory, Al-Muthanna Health Directorate | Department of Virology, Faculty of Medicine, University of Helsinki, Helsinki, Finlan | Nihad Al-Rashedi, Hussein Alburkat, Murad Munahi, Alaa Hameed, Ali Jasim, Olli Vapalahti, Tarja Sironen,Teemu Smura |
| **hCoV-19/Iraq/Samawa-7/2021** | EPI_ISL_1524336 | Biology Department, College of Science, Al Muthanna University and Public Health Laboratory, Al-Muthanna Health Directorate | Department of Virology, Faculty of Medicine, University of Helsinki, Helsinki, Finlan | Nihad Al-Rashedi, Hussein Alburkat, Murad Munahi, Alaa Hameed, Ali Jasim, Olli Vapalahti, Tarja Sironen,Teemu Smura |
| **hCoV-19/Iraq/Samawa-8/2021** | EPI_ISL_1524337 | Biology Department, College of Science, Al Muthanna University and Public Health Laboratory, Al-Muthanna Health Directorate | Department of Virology, Faculty of Medicine, University of Helsinki, Helsinki, Finlan | Nihad Al-Rashedi, Hussein Alburkat, Murad Munahi, Alaa Hameed, Ali Jasim, Olli Vapalahti, Tarja Sironen,Teemu Smura |
| **hCoV-19/Iraq/Samawa-9/2021** | EPI_ISL_1524338 | Biology Department, College of Science, Al Muthanna University and Public Health Laboratory, Al-Muthanna Health Directorate | Department of Virology, Faculty of Medicine, University of Helsinki, Helsinki, Finlan | Nihad Al-Rashedi, Hussein Alburkat, Murad Munahi, Alaa Hameed, Ali Jasim, Olli Vapalahti, Tarja Sironen,Teemu Smura |
| **hCoV-19/Iraq/Samawa-10/2021** | EPI_ISL_1524339 | Biology Department, College of Science, Al Muthanna University and Public Health Laboratory, Al-Muthanna Health Directorate | Department of Virology, Faculty of Medicine, University of Helsinki, Helsinki, Finlan | Nihad Al-Rashedi, Hussein Alburkat, Murad Munahi, Alaa Hameed, Ali Jasim, Olli Vapalahti, Tarja Sironen,Teemu Smura |
| **hCoV-19/Iraq/Samawa-11/2021** | EPI_ISL_1524340 | Biology Department, College of Science, Al Muthanna University and Public Health Laboratory, Al-Muthanna Health Directorate | Department of Virology, Faculty of Medicine, University of Helsinki, Helsinki, Finlan | Nihad Al-Rashedi, Hussein Alburkat, Murad Munahi, Alaa Hameed, Ali Jasim, Olli Vapalahti, Tarja Sironen,Teemu Smura |
| **hCoV-19/Iraq/Samawa-12/2021** | EPI_ISL_1524341 | Biology Department, College of Science, Al Muthanna University and Public Health Laboratory, Al-Muthanna Health Directorate | Department of Virology, Faculty of Medicine, University of Helsinki, Helsinki, Finlan | Nihad Al-Rashedi, Hussein Alburkat, Murad Munahi, Alaa Hameed, Ali Jasim, Olli Vapalahti, Tarja Sironen,Teemu Smura |

| | | | | |
|---|---|---|---|---|
| **hCoV-19/Iraq/Samawa-13/2021** | EPI_ISL_1524342 | Biology Department, College of Science, Al Muthanna University and Public Health Laboratory, Al-Muthanna Health Directorate | Department of Virology, Faculty of Medicine, University of Helsinki, Helsinki, Finlan | Nihad Al-Rashedi, Hussein Alburkat, Murad Munahi, Alaa Hameed, Ali Jasim, Olli Vapalahti, Tarja Sironen,Teemu Smura |
| **hCoV-19/Iraq/Samawa-14/2021** | EPI_ISL_1524343 | Biology Department, College of Science, Al Muthanna University and Public Health Laboratory, Al-Muthanna Health Directorate | Department of Virology, Faculty of Medicine, University of Helsinki, Helsinki, Finlan | Nihad Al-Rashedi, Hussein Alburkat, Murad Munahi, Alaa Hameed, Ali Jasim, Olli Vapalahti, Tarja Sironen,Teemu Smura |
| **hCoV-19/Iraq/Samawa-15/2021** | EPI_ISL_1524344 | Biology Department, College of Science, Al Muthanna University and Public Health Laboratory, Al-Muthanna Health Directorate | Department of Virology, Faculty of Medicine, University of Helsinki, Helsinki, Finlan | Nihad Al-Rashedi, Hussein Alburkat, Murad Munahi, Alaa Hameed, Ali Jasim, Olli Vapalahti, Tarja Sironen,Teemu Smura |
| **hCoV-19/Iraq/Samawa-16/2021** | EPI_ISL_1524345 | Biology Department, College of Science, Al Muthanna University and Public Health Laboratory, Al-Muthanna Health Directorate | Department of Virology, Faculty of Medicine, University of Helsinki, Helsinki, Finlan | Nihad Al-Rashedi, Hussein Alburkat, Murad Munahi, Alaa Hameed, Ali Jasim, Olli Vapalahti, Tarja Sironen,Teemu Smura |
| **hCoV-19/Iraq/Samawa-17/2021** | EPI_ISL_1524346 | Biology Department, College of Science, Al Muthanna University and Public Health Laboratory, Al-Muthanna Health Directorate | Department of Virology, Faculty of Medicine, University of Helsinki, Helsinki, Finlan | Nihad Al-Rashedi, Hussein Alburkat, Murad Munahi, Alaa Hameed, Ali Jasim, Olli Vapalahti, Tarja Sironen,Teemu Smura |
| **hCoV-19/Iraq/Samawa-18/2021** | EPI_ISL_1524347 | Biology Department, College of Science, Al Muthanna University and Public Health Laboratory, Al-Muthanna Health Directorate | Department of Virology, Faculty of Medicine, University of Helsinki, Helsinki, Finlan | Nihad Al-Rashedi, Hussein Alburkat, Murad Munahi, Alaa Hameed, Ali Jasim, Olli Vapalahti, Tarja Sironen,Teemu Smura |
| **hCoV-19/Iraq/Samawa-19/2021** | EPI_ISL_1524348 | Biology Department, College of Science, Al Muthanna University and Public Health Laboratory, Al-Muthanna Health Directorate | Department of Virology, Faculty of Medicine, University of Helsinki, Helsinki, Finlan | Nihad Al-Rashedi, Hussein Alburkat, Murad Munahi, Alaa Hameed, Ali Jasim, Olli Vapalahti, Tarja Sironen,Teemu Smura |
| **hCoV-19/Iraq/Samawa-20/2021** | EPI_ISL_1524349 | Biology Department, College of Science, Al Muthanna University and Public Health Laboratory, Al-Muthanna Health Directorate | Department of Virology, Faculty of Medicine, University of Helsinki, Helsinki, Finlan | Nihad Al-Rashedi, Hussein Alburkat, Murad Munahi, Alaa Hameed, Ali Jasim, Olli Vapalahti, Tarja Sironen,Teemu Smura |
| **hCoV-19/Iraq/Samawa-** | EPI_ISL_1524350 | Biology Department, College of Science, Al Muthanna University and Public Health | Department of Virology, Faculty of Medicine, University of Helsinki, | Nihad Al-Rashedi, Hussein Alburkat, Murad Munahi, Alaa Hameed, Ali Jasim, Olli |

| | | | | |
|---|---|---|---|---|
| **21/2021** | | Laboratory, Al-Muthanna Health Directorate | Helsinki, Finlan | Vapalahti, Tarja Sironen,Teemu Smura |
| **hCoV-19/Iraq/Samawa-22/2021** | EPI_ISL_1524351 | Biology Department, College of Science, Al Muthanna University and Public Health Laboratory, Al-Muthanna Health Directorate | Department of Virology, Faculty of Medicine, University of Helsinki, Helsinki, Finlan | Nihad Al-Rashedi, Hussein Alburkat, Murad Munahi, Alaa Hameed, Ali Jasim, Olli Vapalahti, Tarja Sironen,Teemu Smura |
| **hCoV-19/Iraq/Samawa-23/2021** | EPI_ISL_1524352 | Biology Department, College of Science, Al Muthanna University and Public Health Laboratory, Al-Muthanna Health Directorate | Department of Virology, Faculty of Medicine, University of Helsinki, Helsinki, Finlan | Nihad Al-Rashedi, Hussein Alburkat, Murad Munahi, Alaa Hameed, Ali Jasim, Olli Vapalahti, Tarja Sironen,Teemu Smura |
| **hCoV-19/Iraq/Samawa-24/2021** | EPI_ISL_1524353 | Biology Department, College of Science, Al Muthanna University and Public Health Laboratory, Al-Muthanna Health Directorate | Department of Virology, Faculty of Medicine, University of Helsinki, Helsinki, Finlan | Nihad Al-Rashedi, Hussein Alburkat, Murad Munahi, Alaa Hameed, Ali Jasim, Olli Vapalahti, Tarja Sironen,Teemu Smura |
| **hCoV-19/Iraq/Samawa-25/2021** | EPI_ISL_1524354 | Biology Department, College of Science, Al Muthanna University and Public Health Laboratory, Al-Muthanna Health Directorate | Department of Virology, Faculty of Medicine, University of Helsinki, Helsinki, Finlan | Nihad Al-Rashedi, Hussein Alburkat, Murad Munahi, Alaa Hameed, Ali Jasim, Olli Vapalahti, Tarja Sironen,Teemu Smura |
| **hCoV-19/Iraq/Samawa-26/2021** | EPI_ISL_1524355 | Biology Department, College of Science, Al Muthanna University and Public Health Laboratory, Al-Muthanna Health Directorate | Department of Virology, Faculty of Medicine, University of Helsinki, Helsinki, Finlan | Nihad Al-Rashedi, Hussein Alburkat, Murad Munahi, Alaa Hameed, Ali Jasim, Olli Vapalahti, Tarja Sironen,Teemu Smura |
| **hCoV-19/Iraq/Samawa-27/2021** | EPI_ISL_1524356 | Biology Department, College of Science, Al Muthanna University and Public Health Laboratory, Al-Muthanna Health Directorate | Department of Virology, Faculty of Medicine, University of Helsinki, Helsinki, Finlan | Nihad Al-Rashedi, Hussein Alburkat, Murad Munahi, Alaa Hameed, Ali Jasim, Olli Vapalahti, Tarja Sironen,Teemu Smura |
| **hCoV-19/Iraq/Samawa-28/2021** | EPI_ISL_1524357 | Biology Department, College of Science, Al Muthanna University and Public Health Laboratory, Al-Muthanna Health Directorate | Department of Virology, Faculty of Medicine, University of Helsinki, Helsinki, Finlan | Nihad Al-Rashedi, Hussein Alburkat, Murad Munahi, Alaa Hameed, Ali Jasim, Olli Vapalahti, Tarja Sironen,Teemu Smura |
| **hCoV-19/Iraq/Samawa-29/2021** | EPI_ISL_1524358 | Biology Department, College of Science, Al Muthanna University and Public Health Laboratory, Al-Muthanna Health Directorate | Department of Virology, Faculty of Medicine, University of Helsinki, Helsinki, Finlan | Nihad Al-Rashedi, Hussein Alburkat, Murad Munahi, Alaa Hameed, Ali Jasim, Olli Vapalahti, Tarja Sironen,Teemu Smura |

| | | | | |
|---|---|---|---|---|
| **hCoV-19/Iraq/Samawa-30/2021** | EPI_ISL_1524359 | Biology Department, College of Science, Al Muthanna University and Public Health Laboratory, Al-Muthanna Health Directorate | Department of Virology, Faculty of Medicine, University of Helsinki, Helsinki, Finlan | Nihad Al-Rashedi, Hussein Alburkat, Murad Munahi, Alaa Hameed, Ali Jasim, Olli Vapalahti, Tarja Sironen,Teemu Smura |
| **hCoV-19/Iraq/Samawa-31/2021** | EPI_ISL_1524360 | Biology Department, College of Science, Al Muthanna University and Public Health Laboratory, Al-Muthanna Health Directorate | Department of Virology, Faculty of Medicine, University of Helsinki, Helsinki, Finlan | Nihad Al-Rashedi, Hussein Alburkat, Murad Munahi, Alaa Hameed, Ali Jasim, Olli Vapalahti, Tarja Sironen,Teemu Smura |
| **hCoV-19/Iraq/Samawa-32/2021** | EPI_ISL_1524361 | Biology Department, College of Science, Al Muthanna University and Public Health Laboratory, Al-Muthanna Health Directorate | Department of Virology, Faculty of Medicine, University of Helsinki, Helsinki, Finlan | Nihad Al-Rashedi, Hussein Alburkat, Murad Munahi, Alaa Hameed, Ali Jasim, Olli Vapalahti, Tarja Sironen,Teemu Smura |
| **hCoV-19/Iraq/Samawa-33/2021** | EPI_ISL_1524362 | Biology Department, College of Science, Al Muthanna University and Public Health Laboratory, Al-Muthanna Health Directorate | Department of Virology, Faculty of Medicine, University of Helsinki, Helsinki, Finlan | Nihad Al-Rashedi, Hussein Alburkat, Murad Munahi, Alaa Hameed, Ali Jasim, Olli Vapalahti, Tarja Sironen,Teemu Smura |
| **hCoV-19/Iraq/Samawa-34/2021** | EPI_ISL_1524363 | Biology Department, College of Science, Al Muthanna University and Public Health Laboratory, Al-Muthanna Health Directorate | Department of Virology, Faculty of Medicine, University of Helsinki, Helsinki, Finlan | Nihad Al-Rashedi, Hussein Alburkat, Murad Munahi, Alaa Hameed, Ali Jasim, Olli Vapalahti, Tarja Sironen,Teemu Smura |
| **hCoV-19/Iraq/Samawa-35/2021** | EPI_ISL_1524364 | Biology Department, College of Science, Al Muthanna University and Public Health Laboratory, Al-Muthanna Health Directorate | Department of Virology, Faculty of Medicine, University of Helsinki, Helsinki, Finlan | Nihad Al-Rashedi, Hussein Alburkat, Murad Munahi, Alaa Hameed, Ali Jasim, Olli Vapalahti, Tarja Sironen,Teemu Smura |
| **hCoV-19/Iraq/Samawa-36/2021** | EPI_ISL_1524365 | Biology Department, College of Science, Al Muthanna University and Public Health Laboratory, Al-Muthanna Health Directorate | Department of Virology, Faculty of Medicine, University of Helsinki, Helsinki, Finlan | Nihad Al-Rashedi, Hussein Alburkat, Murad Munahi, Alaa Hameed, Ali Jasim, Olli Vapalahti, Tarja Sironen,Teemu Smura |
| **hCoV-19/Iraq/Samawa-37/2021** | EPI_ISL_1524366 | Biology Department, College of Science, Al Muthanna University and Public Health Laboratory, Al-Muthanna Health Directorate | Department of Virology, Faculty of Medicine, University of Helsinki, Helsinki, Finlan | Nihad Al-Rashedi, Hussein Alburkat, Murad Munahi, Alaa Hameed, Ali Jasim, Olli Vapalahti, Tarja Sironen,Teemu Smura |
| **hCoV-19/Iraq/Samawa-** | EPI_ISL_1524367 | Biology Department, College of Science, Al Muthanna University and Public Health | Department of Virology, Faculty of Medicine, University of Helsinki, | Nihad Al-Rashedi, Hussein Alburkat, Murad Munahi, Alaa Hameed, Ali Jasim, Olli |

| | | | | |
|---|---|---|---|---|
| **38/2021** | | Laboratory, Al-Muthanna Health Directorate | Helsinki, Finlan | Vapalahti, Tarja Sironen,Teemu Smura |
| **hCoV-19/Iraq/Samawa-39/2021** | EPI_ISL_1524368 | Biology Department, College of Science, Al Muthanna University and Public Health Laboratory, Al-Muthanna Health Directorate | Department of Virology, Faculty of Medicine, University of Helsinki, Helsinki, Finlan | Nihad Al-Rashedi, Hussein Alburkat, Murad Munahi, Alaa Hameed, Ali Jasim, Olli Vapalahti, Tarja Sironen,Teemu Smura |
| **hCoV-19/Iraq/Samawa-40/2021** | EPI_ISL_1524369 | Biology Department, College of Science, Al Muthanna University and Public Health Laboratory, Al-Muthanna Health Directorate | Department of Virology, Faculty of Medicine, University of Helsinki, Helsinki, Finlan | Nihad Al-Rashedi, Hussein Alburkat, Murad Munahi, Alaa Hameed, Ali Jasim, Olli Vapalahti, Tarja Sironen,Teemu Smura |
| **hCoV-19/Iraq/Samawa-41/2021** | EPI_ISL_1524370 | Biology Department, College of Science, Al Muthanna University and Public Health Laboratory, Al-Muthanna Health Directorate | Department of Virology, Faculty of Medicine, University of Helsinki, Helsinki, Finlan | Nihad Al-Rashedi, Hussein Alburkat, Murad Munahi, Alaa Hameed, Ali Jasim, Olli Vapalahti, Tarja Sironen,Teemu Smura |
| **hCoV-19/Iraq/Samawa-42/2021** | EPI_ISL_1524371 | Biology Department, College of Science, Al Muthanna University and Public Health Laboratory, Al-Muthanna Health Directorate | Department of Virology, Faculty of Medicine, University of Helsinki, Helsinki, Finlan | Nihad Al-Rashedi, Hussein Alburkat, Murad Munahi, Alaa Hameed, Ali Jasim, Olli Vapalahti, Tarja Sironen,Teemu Smura |
| **hCoV-19/Iraq/Samawa-43/2021** | EPI_ISL_1524372 | Biology Department, College of Science, Al Muthanna University and Public Health Laboratory, Al-Muthanna Health Directorate | Department of Virology, Faculty of Medicine, University of Helsinki, Helsinki, Finlan | Nihad Al-Rashedi, Hussein Alburkat, Murad Munahi, Alaa Hameed, Ali Jasim, Olli Vapalahti, Tarja Sironen,Teemu Smura |
| **hCoV-19/Iraq/Samawa-44/2021** | EPI_ISL_1524373 | Biology Department, College of Science, Al Muthanna University and Public Health Laboratory, Al-Muthanna Health Directorate | Department of Virology, Faculty of Medicine, University of Helsinki, Helsinki, Finlan | Nihad Al-Rashedi, Hussein Alburkat, Murad Munahi, Alaa Hameed, Ali Jasim, Olli Vapalahti, Tarja Sironen,Teemu Smura |
| **hCoV-19/Iraq/Samawa-45/2021** | EPI_ISL_1524374 | Biology Department, College of Science, Al Muthanna University and Public Health Laboratory, Al-Muthanna Health Directorate | Department of Virology, Faculty of Medicine, University of Helsinki, Helsinki, Finlan | Nihad Al-Rashedi, Hussein Alburkat, Murad Munahi, Alaa Hameed, Ali Jasim, Olli Vapalahti, Tarja Sironen,Teemu Smura |
| **hCoV-19/Iraq/Samawa-46/2021** | EPI_ISL_1524375 | Biology Department, College of Science, Al Muthanna University and Public Health Laboratory, Al-Muthanna Health Directorate | Department of Virology, Faculty of Medicine, University of Helsinki, Helsinki, Finlan | Nihad Al-Rashedi, Hussein Alburkat, Murad Munahi, Alaa Hameed, Ali Jasim, Olli Vapalahti, Tarja Sironen,Teemu Smura |

| | | | | |
|---|---|---|---|---|
| **hCoV-19/Iraq/Samawa-47/2021** | EPI_ISL_1524376 | Biology Department, College of Science, Al Muthanna University and Public Health Laboratory, Al-Muthanna Health Directorate | Department of Virology, Faculty of Medicine, University of Helsinki, Helsinki, Finlan | Nihad Al-Rashedi, Hussein Alburkat, Murad Munahi, Alaa Hameed, Ali Jasim, Olli Vapalahti, Tarja Sironen,Teemu Smura |
| **hCoV-19/Iraq/Samawa-48/2021** | EPI_ISL_1524377 | Biology Department, College of Science, Al Muthanna University and Public Health Laboratory, Al-Muthanna Health Directorate | Department of Virology, Faculty of Medicine, University of Helsinki, Helsinki, Finlan | Nihad Al-Rashedi, Hussein Alburkat, Murad Munahi, Alaa Hameed, Ali Jasim, Olli Vapalahti, Tarja Sironen,Teemu Smura |
| **hCoV-19/Iraq/Samawa-49/2021** | EPI_ISL_1524378 | Biology Department, College of Science, Al Muthanna University and Public Health Laboratory, Al-Muthanna Health Directorate | Department of Virology, Faculty of Medicine, University of Helsinki, Helsinki, Finlan | Nihad Al-Rashedi, Hussein Alburkat, Murad Munahi, Alaa Hameed, Ali Jasim, Olli Vapalahti, Tarja Sironen,Teemu Smura |
| **hCoV-19/Iraq/Samawa-50/2021** | EPI_ISL_1524379 | Biology Department, College of Science, Al Muthanna University and Public Health Laboratory, Al-Muthanna Health Directorate | Department of Virology, Faculty of Medicine, University of Helsinki, Helsinki, Finlan | Nihad Al-Rashedi, Hussein Alburkat, Murad Munahi, Alaa Hameed, Ali Jasim, Olli Vapalahti, Tarja Sironen,Teemu Smura |
| **hCoV-19/Iraq/Samawa-51/2021** | EPI_ISL_1524380 | Biology Department, College of Science, Al Muthanna University and Public Health Laboratory, Al-Muthanna Health Directorate | Department of Virology, Faculty of Medicine, University of Helsinki, Helsinki, Finlan | Nihad Al-Rashedi, Hussein Alburkat, Murad Munahi, Alaa Hameed, Ali Jasim, Olli Vapalahti, Tarja Sironen,Teemu Smura |
| **hCoV-19/Iraq/Samawa-52/2021** | EPI_ISL_1524381 | Biology Department, College of Science, Al Muthanna University and Public Health Laboratory, Al-Muthanna Health Directorate | Department of Virology, Faculty of Medicine, University of Helsinki, Helsinki, Finlan | Nihad Al-Rashedi, Hussein Alburkat, Murad Munahi, Alaa Hameed, Ali Jasim, Olli Vapalahti, Tarja Sironen,Teemu Smura |
| **hCoV-19/Iraq/Samawa-53/2021** | EPI_ISL_2467906 | Biology Department, College of Science, Al Muthanna University and Public Health Laboratory, Al-Muthanna Health Directorate | Department of Virology, Faculty of Medicine, University of Helsinki, Helsinki, Finlan | Nihad Al-Rashedi, Hussein Alburkat, Murad Munahi, Alaa Hameed, Ali Jasim, Olli Vapalahti, Tarja Sironen,Teemu Smura |
| **hCoV-19/Iraq/Samawa-54/2021** | EPI_ISL_2467907 | Biology Department, College of Science, Al Muthanna University and Public Health Laboratory, Al-Muthanna Health Directorate | Department of Virology, Faculty of Medicine, University of Helsinki, Helsinki, Finlan | Nihad Al-Rashedi, Hussein Alburkat, Murad Munahi, Alaa Hameed, Ali Jasim, Olli Vapalahti, Tarja Sironen,Teemu Smura |
| **hCoV-19/Iraq/Samawa-** | EPI_ISL_2467908 | Biology Department, College of Science, Al Muthanna University and Public Health | Department of Virology, Faculty of Medicine, University of Helsinki, | Nihad Al-Rashedi, Hussein Alburkat, Murad Munahi, Alaa Hameed, Ali Jasim, Olli |

| | | | | |
|---|---|---|---|---|
| **55/2021** | | Laboratory, Al-Muthanna Health Directorate | Helsinki, Finlan | Vapalahti, Tarja Sironen,Teemu Smura |
| **hCoV-19/Iraq/Samawa-56/2021** | EPI_ISL_2467909 | Biology Department, College of Science, Al Muthanna University and Public Health Laboratory, Al-Muthanna Health Directorate | Department of Virology, Faculty of Medicine, University of Helsinki, Helsinki, Finlan | Nihad Al-Rashedi, Hussein Alburkat, Murad Munahi, Alaa Hameed, Ali Jasim, Olli Vapalahti, Tarja Sironen,Teemu Smura |
| **hCoV-19/Iraq/Samawa-57/2021** | EPI_ISL_2467910 | Biology Department, College of Science, Al Muthanna University and Public Health Laboratory, Al-Muthanna Health Directorate | Department of Virology, Faculty of Medicine, University of Helsinki, Helsinki, Finlan | Nihad Al-Rashedi, Hussein Alburkat, Murad Munahi, Alaa Hameed, Ali Jasim, Olli Vapalahti, Tarja Sironen,Teemu Smura |
| **hCoV-19/Iraq/Samawa-58/2021** | EPI_ISL_2467911 | Biology Department, College of Science, Al Muthanna University and Public Health Laboratory, Al-Muthanna Health Directorate | Department of Virology, Faculty of Medicine, University of Helsinki, Helsinki, Finlan | Nihad Al-Rashedi, Hussein Alburkat, Murad Munahi, Alaa Hameed, Ali Jasim, Olli Vapalahti, Tarja Sironen,Teemu Smura |
| **hCoV-19/Iraq/Samawa-59/2021** | EPI_ISL_2467912 | Biology Department, College of Science, Al Muthanna University and Public Health Laboratory, Al-Muthanna Health Directorate | Department of Virology, Faculty of Medicine, University of Helsinki, Helsinki, Finlan | Nihad Al-Rashedi, Hussein Alburkat, Murad Munahi, Alaa Hameed, Ali Jasim, Olli Vapalahti, Tarja Sironen,Teemu Smura |
| **hCoV-19/Iraq/Samawa-60/2021** | EPI_ISL_2467913 | Biology Department, College of Science, Al Muthanna University and Public Health Laboratory, Al-Muthanna Health Directorate | Department of Virology, Faculty of Medicine, University of Helsinki, Helsinki, Finlan | Nihad Al-Rashedi, Hussein Alburkat, Murad Munahi, Alaa Hameed, Ali Jasim, Olli Vapalahti, Tarja Sironen,Teemu Smura |
| **hCoV-19/Iraq/Samawa-61/2021** | EPI_ISL_2467914 | Biology Department, College of Science, Al Muthanna University and Public Health Laboratory, Al-Muthanna Health Directorate | Department of Virology, Faculty of Medicine, University of Helsinki, Helsinki, Finlan | Nihad Al-Rashedi, Hussein Alburkat, Murad Munahi, Alaa Hameed, Ali Jasim, Olli Vapalahti, Tarja Sironen,Teemu Smura |
| **hCoV-19/Iraq/Samawa-62/2021** | EPI_ISL_2467915 | Biology Department, College of Science, Al Muthanna University and Public Health Laboratory, Al-Muthanna Health Directorate | Department of Virology, Faculty of Medicine, University of Helsinki, Helsinki, Finlan | Nihad Al-Rashedi, Hussein Alburkat, Murad Munahi, Alaa Hameed, Ali Jasim, Olli Vapalahti, Tarja Sironen,Teemu Smura |
| **hCoV-19/Iraq/Samawa-63/2021** | EPI_ISL_2467916 | Biology Department, College of Science, Al Muthanna University and Public Health Laboratory, Al-Muthanna Health Directorate | Department of Virology, Faculty of Medicine, University of Helsinki, Helsinki, Finlan | Nihad Al-Rashedi, Hussein Alburkat, Murad Munahi, Alaa Hameed, Ali Jasim, Olli Vapalahti, Tarja Sironen,Teemu Smura |

| | | | | |
|---|---|---|---|---|
| **hCoV-19/Iraq/Samawa-64/2021** | EPI_ISL_2467917 | Biology Department, College of Science, Al Muthanna University and Public Health Laboratory, Al-Muthanna Health Directorate | Department of Virology, Faculty of Medicine, University of Helsinki, Helsinki, Finlan | Nihad Al-Rashedi, Hussein Alburkat, Murad Munahi, Alaa Hameed, Ali Jasim, Olli Vapalahti, Tarja Sironen,Teemu Smura |
| **hCoV-19/Iraq/Samawa-65/2021** | EPI_ISL_2467918 | Biology Department, College of Science, Al Muthanna University and Public Health Laboratory, Al-Muthanna Health Directorate | Department of Virology, Faculty of Medicine, University of Helsinki, Helsinki, Finlan | Nihad Al-Rashedi, Hussein Alburkat, Murad Munahi, Alaa Hameed, Ali Jasim, Olli Vapalahti, Tarja Sironen,Teemu Smura |
| **hCoV-19/Iraq/Samawa-66/2021** | EPI_ISL_2467919 | Biology Department, College of Science, Al Muthanna University and Public Health Laboratory, Al-Muthanna Health Directorate | Department of Virology, Faculty of Medicine, University of Helsinki, Helsinki, Finlan | Nihad Al-Rashedi, Hussein Alburkat, Murad Munahi, Alaa Hameed, Ali Jasim, Olli Vapalahti, Tarja Sironen,Teemu Smura |
| **hCoV-19/Iraq/Samawa-67/2021** | EPI_ISL_2467920 | Biology Department, College of Science, Al Muthanna University and Public Health Laboratory, Al-Muthanna Health Directorate | Department of Virology, Faculty of Medicine, University of Helsinki, Helsinki, Finlan | Nihad Al-Rashedi, Hussein Alburkat, Murad Munahi, Alaa Hameed, Ali Jasim, Olli Vapalahti, Tarja Sironen,Teemu Smura |
| **hCoV-19/Iraq/Samawa-69/2021** | EPI_ISL_2467921 | Biology Department, College of Science, Al Muthanna University and Public Health Laboratory, Al-Muthanna Health Directorate | Department of Virology, Faculty of Medicine, University of Helsinki, Helsinki, Finlan | Nihad Al-Rashedi, Hussein Alburkat, Murad Munahi, Alaa Hameed, Ali Jasim, Olli Vapalahti, Tarja Sironen,Teemu Smura |
| **hCoV-19/Iraq/Samawa-71/2021** | EPI_ISL_2467922 | Biology Department, College of Science, Al Muthanna University and Public Health Laboratory, Al-Muthanna Health Directorate | Department of Virology, Faculty of Medicine, University of Helsinki, Helsinki, Finlan | Nihad Al-Rashedi, Hussein Alburkat, Murad Munahi, Alaa Hameed, Ali Jasim, Olli Vapalahti, Tarja Sironen,Teemu Smura |
| **hCoV-19/Iraq/Samawa-72/2021** | EPI_ISL_2467923 | Biology Department, College of Science, Al Muthanna University and Public Health Laboratory, Al-Muthanna Health Directorate | Department of Virology, Faculty of Medicine, University of Helsinki, Helsinki, Finlan | Nihad Al-Rashedi, Hussein Alburkat, Murad Munahi, Alaa Hameed, Ali Jasim, Olli Vapalahti, Tarja Sironen,Teemu Smura |
| **hCoV-19/Iraq/Samawa-73/2021** | EPI_ISL_2467924 | Biology Department, College of Science, Al Muthanna University and Public Health Laboratory, Al-Muthanna Health Directorate | Department of Virology, Faculty of Medicine, University of Helsinki, Helsinki, Finlan | Nihad Al-Rashedi, Hussein Alburkat, Murad Munahi, Alaa Hameed, Ali Jasim, Olli Vapalahti, Tarja Sironen,Teemu Smura |
| **hCoV-19/Iraq/Samawa-** | EPI_ISL_2467925 | Biology Department, College of Science, Al Muthanna University and Public Health | Department of Virology, Faculty of Medicine, University of Helsinki, | Nihad Al-Rashedi, Hussein Alburkat, Murad Munahi, Alaa Hameed, Ali Jasim, Olli |

| | | | | |
|---|---|---|---|---|
| **74/2021** | | Laboratory, Al-Muthanna Health Directorate | Helsinki, Finlan | Vapalahti, Tarja Sironen,Teemu Smura |
| **hCoV-19/Iraq/Samawa-75/2021** | EPI_ISL_2467926 | Biology Department, College of Science, Al Muthanna University and Public Health Laboratory, Al-Muthanna Health Directorate | Department of Virology, Faculty of Medicine, University of Helsinki, Helsinki, Finlan | Nihad Al-Rashedi, Hussein Alburkat, Murad Munahi, Alaa Hameed, Ali Jasim, Olli Vapalahti, Tarja Sironen,Teemu Smura |
| **hCoV-19/Iraq/Samawa-76/2021** | EPI_ISL_2467927 | Biology Department, College of Science, Al Muthanna University and Public Health Laboratory, Al-Muthanna Health Directorate | Department of Virology, Faculty of Medicine, University of Helsinki, Helsinki, Finlan | Nihad Al-Rashedi, Hussein Alburkat, Murad Munahi, Alaa Hameed, Ali Jasim, Olli Vapalahti, Tarja Sironen,Teemu Smura |
| **hCoV-19/Iraq/Samawa-77/2021** | EPI_ISL_2467928 | Biology Department, College of Science, Al Muthanna University and Public Health Laboratory, Al-Muthanna Health Directorate | Department of Virology, Faculty of Medicine, University of Helsinki, Helsinki, Finlan | Nihad Al-Rashedi, Hussein Alburkat, Murad Munahi, Alaa Hameed, Ali Jasim, Olli Vapalahti, Tarja Sironen,Teemu Smura |
| **hCoV-19/Iraq/Samawa-78/2021** | EPI_ISL_2467929 | Biology Department, College of Science, Al Muthanna University and Public Health Laboratory, Al-Muthanna Health Directorate | Department of Virology, Faculty of Medicine, University of Helsinki, Helsinki, Finlan | Nihad Al-Rashedi, Hussein Alburkat, Murad Munahi, Alaa Hameed, Ali Jasim, Olli Vapalahti, Tarja Sironen,Teemu Smura |
| **hCoV-19/Oman/RESP-20-4400/2020** | EPI_ISL_457990 | Oman-NIC | Oman-NIC | Samira Al-Maruqi, Fahad Zadjali, Amina Al Jardani, Khulood Al-Mammary, Hanan Al-kindi, Fatma BaAlawi, Hamida AL Barwani, Zeyana AL-Dahmani, Intisar Al-Shukri, Aisha Al-Busaidi, Aisha Al-Amri, Ahlam Al-Amri, Mohammed Al-Tobi, Samiha Al Kharusi, Abdulla Balkhair |
| **hCoV-19/Senegal/306/2020** | EPI_ISL_420069 | Institut Pasteur Dakar | Institut Pasteur de Dakar | Ndongo Dia, Moussa Moise Diagne, Mamadou Diop, Ousmane Faye, Amadou Alpha Sall<br><br>Submitter information |
| **hCoV-19/Peru/LAM-UPCH-0006/2020** | EPI_ISL_529067 | Laboratorio de Referencia Nacional de Virus Respiratorios, Instituto Nacional de Salud | Laboratorio de Genómica Microbiana, Universidad Peruana | Pablo Tsukayama, Alejandra Dávila-Barclay, Luis González, Pedro E. Romero, Brenda Ayzanoa, Janet Huancachoque, Pool |

| | | | | |
|---|---|---|---|---|
| | | Peru | Cayetano Heredia | Marcos, Maribel Huaringa |
| **hCoV-19/Peru/LIM-INS-333/2020** | EPI_ISL_1111231 | Laboratorio de Referencia Nacional de Virus Respiratorio. Instituto Nacional de Salud Perú | Laboratorio de Referencia Nacional de Enteropatógenos. Instituto Nacional de Salud del Perú | Ronnie Gavilan Chavez, Junior Caro Castro, Willi Quino Sifuentes, Veronica Hurtado Vela, Iris Silva Molina, Fiorella Orellana Peralta |
| **hCoV-19/Beijing/DT-BJ04/2020** | EPI_ISL_452364 | Laboratory of Infectious Diseases Center of Beijing Ditan Hospital | Laboratory of Infectious Diseases Center of Beijing Ditan Hospital | Siyuan Yang, Chengjie Jie, Fengting Yu, Yunxia Tang, Liting Yan, Linghang Wang |
| **hCoV-19/Argentina/PAIS-A0222/2020** | EPI_ISL_792303 | Unidad de Virología, Centro de Educación Médica en Investigaciones Clínicas CEMIC | Área de Secuenciación del Laboratorio de Virología del Hospital de Niños Dr. Ricardo Gutierrez on behalf of 'Proyecto Argentino Interinstitucional de genomica de SARS-CoV-2' (PAIS Consortium) | Nabaes Jodar, MS; Goya, S; Natale, MI; Lusso, S; Echavarría, M; Mistchenko, AS; Valinotto, LE; Viegas, M. |
| **hCoV-19/Algeria/G35014-8856/2020** | EPI_ISL_766869 | NIC Viral Respiratory Unit - Institut Pasteur of Algeria | National Reference Center for Viruses of Respiratory Infections, Institut Pasteur, Paris | Mélanie Albert, Marion Barbet, Sylvie Behillil, Méline Bizard, Angela Brisebarre, Flora Donati, Etienne Simon-Lorière, Vincent Enouf, Maud Vanpeene, Sylvie van der Werf, Fawzi Derrar |
| **hCoV-19/Denmark/ALAB-SSI450/2020** | EPI_ISL_429537 | Department of Virus and Microbiological Special Diagnostics, Statens Serum Institut, Copenhagen, Denmark, Artillerivej 5, 2300 Copenahgen S | Albertsen lab, Department of Chemistry and Bioscience, Aalborg University, Denmark | Rasmus Kirkegaard |
| **hCoV-19/Belgium/DJMG-0325156/2020** | EPI_ISL_420413 | KU Leuven, Clinical and Epidemiological Virology | KU Leuven, Clinical and Epidemiological Virology | Joan Marti-Carreras, Bert Vanmechelen, Tony Wawina, Piet Maes |
| **hCoV-19/Denmark/ALAB-** | EPI_ISL_451989 | Department of Clinical Microbiology, Copenhagen University Hospital, Hvidovre, | Albertsen lab, Department of Chemistry and Bioscience, Aalborg | Rasmus Kirkegaard |

| | | | | |
|---|---|---|---|---|
| **HH-102/2020** | | Kettegaard Alle 30, 2650 Hvidovre | University, Denmark | |
| **hCoV-19/Scotland/EDB075/2020** | EPI_ISL_425872 | Virology Department, Royal Infirmary of Edinburgh, NHS Lothian / School of Biological Sciences, University of Edinburgh / Institute of Genetics and Molecular Medicine, University of Edinburgh | COVID-19 Genomics UK (COG-UK) Consortium | McHugh M, Dewar R, Rooke S, Gallagher M, Balcaza C, O'Toole A, Hill V, McCrone JT, Colquhoun R, Yu X, Jackson B, Scher E, Rambaut A, Williams TC, Templeton K |
| **hCoV-19/Algeria/G37318-8849/2020** | EPI_ISL_766863 | NIC Viral Respiratory Unit - Institut Pasteur of Algeria | National Reference Center for Viruses of Respiratory Infections, Institut Pasteur, Paris | Mélanie Albert, Marion Barbet, Sylvie Behillil, Méline Bizard, Angela Brisebarre, Flora Donati, Etienne Simon-Lorière, Vincent Enouf, Maud Vanpeene, Sylvie van der Werf, Fawzi Derrar |
| **hCoV-19/Canada/ON-PHL-21-16162/2021** | EPI_ISL_2252203 | Public Health Ontario Laboratory | Public Health Ontario Laboratory | Vanessa G Allen, Philip Banh, Yao Chen, Richard de Borja, Alireza Eshaghi, Nahuel Fittipaldi, Christine Frantz, Jonathan B Gubbay, Jennifer L Guthrie, Lawrence Heisler, Esha Joshi, Michael Laszloffy, Aimin Li, Michael CY Li, Dean Maxwell, Sandeep Nagra, Samir N Patel, Jared Simpson, Karthikeyan Sivaraman, Ashleigh Sullivan, Yogi Sundaravadanam, Sarah Teatero, Andre Villegas, Matthew Watson, Sandra Zittermann |
| **hCoV-19/India/GJ-ICMR-NIV-INSACOG-GSEQ-94/2021** | EPI_ISL_1703906 | ICMR-National Institute of Virology - INSACOG | NIV Influenza | Dr. Varsha Potdar |
| **hCoV-19/Germany/NW-RKI-I-081793/2021** | EPI_ISL_1646809 | SYNLAB MVZ Leverkusen | Robert Koch Institute | Drechsel, Oliver |

| | | | | |
|---|---|---|---|---|
| **hCoV-19/Argentina/INEI101331/2021** | EPI_ISL_2135137 | Servicio Virosis Respiratorias-Departamento Virología-INEI | Instituto Nacional Enfermedades Infecciosas C.G.Malbran | Baumeister E., Avaro M., Benedetti E., Russo M., Dattero ME, Pontoriero A., Cisterna D., Molina V., Perandones C., Tuduri E., Lorenzo F., Poklepovich T., Campos J. |
| **hCoV-19/Jordan/Biolab0063/2021** | EPI_ISL_2617455 | | | [illegible] Tibi, Lama [illegible]a Saddedin, Eiad Atwa, Amid Abdelnour |
| **hCoV-19/Iraq/Erbil 5/2021** | EPI_ISL_2629240 | Biology Department, Salahaddin University | Biology Department, Salahaddin University | Khailany,R.A., Rahman,M.O., Ozaslan,M., Faraidun,H.N., Ibrahim,O.Q., Hama,H.A. and Kanabe,B.O. |
| **hCoV-19-Iraq/Erbil 4/2021** | EPI_ISL_2234383 | Biology Department, Salahaddin University | Biology Department, Salahaddin University | Khailany,R.A., Rahman,M.O., Ozaslan,M., Faraidun,H.N., Ibrahim,O.Q., Hama,H.A. and Kanabe,B.O. |
| **hCoV-19/env/Liechtenstein/CeMM8894/2021** | EPI_ISL_2232965 | Department of Microbiology, University Innsbruck | Bergthaler laboratory, CeMM Research Center for Molecular Medicine of the Austrian Academy of Sciences | Lukas Endler, Anna Schedl, Fabian Amman, Petr Triska, Thomas Penz, Benedikt Agerer, Maelle Le Moing, Michael Schuster, Bekir Erguner, Jan Laine, Martin Senekowitsch, Christoph Bock, Andreas Bergthaler |
| **hCoV-19/Germany/SN-RKI-I-024907/2021** | EPI_ISL_1155081 | Bioscientia Labor Wermsdorf | Robert Koch Institute | Drechsel, Oliver |
| **hCoV-19/Germany/BB-RKI-I-026259/2021** | EPI_ISL_1156444 | LADR MVZ Zweigpraxis Bernau | IMS-10036-CVDP-CAC2918B-8F48-491E-BD08-27ADDBE45777 | Drechsel, Oliver |
| **hCoV-19/France/IDF-CERBAHC-20211020-23/2021** | EPI_ISL_862044 | CERBA | CERBA LAB | Roquebert B; Costa JM; Hedbaut E; Trombert S; Lecorche E; Verdurme L; Malek Ramdane, Olivi M; Haïm-Boukobza S. |
| **hCoV-19/Belgium/UGent-** | EPI_ISL_2191955 | Lab voor klinische biologie | Lab voor klinische biologie | Marija Janevska, Hannelore Hamerlinck, Bruno Verhasselt |

| | | | | |
|---|---|---|---|---|
| **5621/2021** | | | | |
| **hCoV-19/Italy/CAM-TIGEM-IZSM-COLLI-17384/2021** | EPI_ISL_3014333 | Università Federico II - Dipartimento di scienze mediche traslazionali - Napoli | TIGEM | Antonio Grimaldi Patrizia Annunziata Francesco Panariello Teresa Giuliano Michele Cennamo Valentina Bouche Chiara Colantuono Lucio Di Filippo Mariano Fiorenza Anna Manfredi Marcello Salvi Giuseppe Portella Andrea Ballabio Davide Cacchiarelli |
| **hCoV-19/Estonia/Cov10119/2021** | EPI_ISL_2643420 | SYNLAB Eesti OÜ | Department of Microbiology, Institute of Biomedicine and Translational Medicine, University of Tartu | Radko Avi, Aare Abroi, Irja Lutsar, Kristi Huik, Taavi Päll, Meri Pauskar, Ene-Ly Jõgeda, Arina Shablinskaja, Eveli Kallas, Kai Truusalu, Dagmar Hoidmets, Katrin Kaarna, Tuuli Reisberg, Lili Azin Milani, Ulvi Gerst Talas, Heiki Niglas, Olga Sadikova, Liidia Dotsenko, Mari-Anne Härma, Kaisa Truus, Mats Hansen, Paul Naaber, Andrio Lahesaare |
| **hCoV-19/England/OXON-F56F69/2021** | EPI_ISL_999154 | Oxford Viromics, NDM, University of Oxford; Oxford University Hospitals; Basingstoke and North Hampshire Hospital | COVID-19 Genomics UK (COG-UK) Consortium | anya Golubchik, David Bonsall, George Macintyre, Amy Trebes, Mariateresa de Cesare, Catrin Moore, Alex Mobbs, Anita Justice, Robert Shaw, Monique Andersson, Timothy Peto, Emma Wise, Nathan Moore, Jessica Lynch, Nick Cortes, Matilde Mori, Stephen Kidd, David Buck, John Todd, Christophe Fraser |
| **hCoV-19/Hong Kong/HKU-210308-Imp4/2020** | EPI_ISL_1197085 | Department of Microbiology, The University of Hong Kong | Department of Microbiology, The University of Hong Kong | Kelvin K.W. To, Kwok-Yung Yuen |
| **hCoV-19/Italy/CAL-AOCatanzaro-05011136/2021** | EPI_ISL_2282248 | Azienda Ospedaliera Pugliese Ciaccio di Catanzaro SOC Microbiologia e Virologia | Azienda Ospedaliera Pugliese Ciaccio di Catanzaro SOC Microbiologia e Virologia | Rossana Tallerico Federica Pasceri Marco De Fazio Ilenia Talotta Giuseppina Panduri Pasquale Minchella |
| **hCoV-19/Switzerland/ZH-UZH-IMV-** | EPI_ISL_1921818 | UniversitŠtsSpital ZŸrich | Institute of Medical Virology | Daniel Ehrsam, Isabel StŸrmer, Catharine Aquino, Joel Wirz, Weihong Qi, Hubert Rehrauer, Verena Kufner, Gabriela Ziltener, |

| | | | | |
|---|---|---|---|---|
| **4555572d/2021** | | | | Maryam Zaheri, Stefan Schmutz, Annette AudigŽ, Maria GrŸnberg, Kevin Steiner, Jon Huder, Cyril Shah, Riccarda Capaul, Guido Bloemberg, JŸrg Bšni, Michael Huber, Alexandra Trkola |
| **hCoV-19/England/OXON-F6044F/2021** | EPI_ISL_999443 | Oxford Viromics, NDM, University of Oxford; Oxford University Hospitals; Basingstoke and North Hampshire Hospital | COVID-19 Genomics UK (COG-UK) Consortium | Tanya Golubchik, David Bonsall, George Macintyre, Amy Trebes, Mariateresa de Cesare, Catrin Moore, Alex Mobbs, Anita Justice, Robert Shaw, Monique Andersson, Timothy Peto, Emma Wise, Nathan Moore, Jessica Lynch, Nick Cortes, Matilde Mori, Stephen Kidd, David Buck, John Todd, Christophe Fraser |
| **hCoV-19/env/Austria/CeMM8358/2021** | EPI_ISL_2137174 | Department of Microbiology, University Innsbruck | Bergthaler laboratory, CeMM Research Center for Molecular Medicine of the Austrian Academy of Sciences | Lukas Endler, Anna Schedl, Fabian Amman, Petr Triska, Thomas Penz, Benedikt Agerer, Maelle Le Moing, Michael Schuster, Bekir Erguner, Jan Laine, Martin Senekowitsch, Christoph Bock, Andreas Bergthaler |
| **hCoV-19/USA/CO-CDPHE-2100874634/2021** | EPI_ISL_2277317 | Colorado Department of Public Health and Environment | Colorado Department of Public Health and Environment | Laura Bankers, Molly C. Hetherington-Rauth, Diana Ir, Alexandria Rossheim, Shannon R. Matzinger, Sarah Elizabeth Totten, Emily A. Travanty |
| **hCoV-19/USA/WI-WSLH-214677/2021** | EPI_ISL_2279344 | Wisconsin State Laboratory of Hygiene Communicable Disease Division | Wisconsin State Laboratory of Hygiene Communicable Disease Division | Abigail C. Shockey, Alicia J. Mooney, Erika M. Hanson, Tonya Danz, Richard Griesser, Sara Wagner, Kelsey R. Florek |
| **hCoV-19/Cameroon/CAM-31/2021** | EPI_ISL_1972314 | Laboratoire National de Santé Publique du Cameroun | Pathogen Genomics Lab, National Institute for Biomedical Research (INRB) | Placide Mbala-Kingebeni, Marie Claire Okomo, Edith Nkwembe, Eddy Kinganda-Lusamaki, Amuri Aziza, Francisca Muyembe Mawete, Emmanuel Lokilo Lofiko, Jean Claude Makangara, Raphael Lumembe, Gabriel Kabamba, Catherine Pratt, Matthias Pauthner, Josh Quick, Trevor Bedford, Ian Goodfellow, Andrew Rambaut, Nick Loman, Michael Wiley, Steve Ahuka-Mundeke, Jean-Jacques Muyembe Tamfum |

| | | | | |
|---|---|---|---|---|
| **hCoV-19/USA TX-DSHS-6592/2021** | EPI_ISL_2280052 | TXDSHS | TXDSHS | Rashmi Tuladhar, Bonnie Oh, Jenny Zhang, Maliha Rahman, Mayela Pedrueza, Anita Pokharel, Karen Bobier, Lorraine Rodriguez, Myong Koag, Chun Wang, Rachel Lee, Grace Kubin |
| **hCoV-19/USA/OH-UHTL-992/2021** | EPI_ISL_2661529 | UHTL, University Hospitals | UHTL, University Hospitals | Sadri,N., Alouani,D., Song,X |
| **hCoV-19/Spain/CM-ISCIII-213286/2021** | EPI_ISL_2331773 | HOSPITAL GENERAL UNIVERSITARIO DE ALBACETE | Instituto de Salud Carlos III | Iglesias-Caballero, M. Sandonís,V. Vázquez-Morón, S. Camarero, S. Pozo, F. Casas, I. Jiménez, P. Zaballos, A. Monzón, S. Varona, S. Cuesta, I.SAINZ DE BARANDA CAMINO, CARIDAD |
| **hCoV-19/Greece/V1183/2021** | EPI_ISL_2235291 | Department of Microbiology, AHEPA University Hospital | Institute of Applied Biosciences, Centre for Research and Technology Hellas | Anastasia Chatzidimitriou et al. |
| **hCoV-19/Albania/210901049/2021** | EPI_ISL_1299858 | Unit of lab surveillance of viral emerging diseases, National Lab of Influenza | Respiratory Virus Unit, National Infection Service, Public Health England | PHE Covid Sequencing Team, Iris Hasibra, Prof Silvia Bino, Prof Albana Fico |
| **hCoV-19/Cambodia/02-2105120711/2021** | EPI_ISL_2231569 | Battambang Provincial Laboratory | Virology Unit, Institut Pasteur du Cambodge | Jurre Y Siegers, Cecile Troupin, Leakhena Pum, Chiek Sivhour, Ly Sovann, Kraing Sidonn, Yi Sengdoeurn, Chin Savuth, Chau Darapheak, Veasna Duong, Erik A Karlsson |
| **hCoV-19/DRC/216884/2021** | EPI_ISL_2135839 | Viral Respiratory Lab, National Institute for Biomedical Research (INRB) | Pathogen Sequencing Lab, National Institute for Biomedical Research (INRB) | Placide Mbala-Kingebeni, Edith Nkwembe, Eddy Kinganda-Lusamaki, Amuri Aziza, Francisca Muyembe Mawete, Emmanuel Lokilo Lofiko, Jean Claude Makangara, Catherine Pratt, Matthias Pauthner, Josh Quick, Allison Black, James Hadfield, Trevor Bedford, Ian Goodfellow, Andrew Rambaut, Nick Loman, Kristian Andersen, Michael Wiley, Steve Ahuka-Mundeke, Jean-Jacques Muyembe Tamfum |
| **hCoV-19/Kuwait/KU-** | EPI_ISL_2604034 | Ministry of Health, Jaber Al-Ahmad Hospital | Virology Unit, Department of | Nada Madi, Hussain Safar, Ebaa Al-Awadhi, Anfal Al-Adwani |

| | | | | |
|---|---|---|---|---|
| **96/2021** | | | Microbiology, Faculty of Medicine, Kuwait University | |
| **hCoV-19/Hong Kong/CM21000226/2021** | EPI_ISL_1963623 | Hong Kong Department of Health | School of Public Health, The University of Hong Kong | Dominic N.C. Tsang, Daniel K.W. Chu, Haogao Gu, Tong Zhang, Leo L.M. Poon, Malik Peiris |
| **hCoV-19/Switzerland/SO-UHB-001-31-551393-61/2021** | EPI_ISL_2858611 | Viollier AG | Clinical Bacteriology | Tim Roloff, Fanny Wegner, Helena MB Seth-Smith, Alfredo Mari, Karoline Leuzinger, Julia Bielicki, Christiane Beckmann, Manuel Battegay, Hans Hirsch, Adrian Egli |
| **hCoV-19/Austria/CeMM7670/2021** | EPI_ISL_1840940 | Austrian Agency for Health and Food Safety (AGES) | Bergthaler laboratory, CeMM Research Center for Molecular Medicine of the Austrian Academy of Sciences | Lukas Endler, Anna Schedl, Fabian Amman, Petr Triska, Thomas Penz, Benedikt Agerer, Maelle Le Moing, Michael Schuster, Bekir Erguner, Jan Laine, Martin Senekowitsch, Christoph Bock, Andreas Bergthaler |
| **hCoV-19/India/PB-ICMR-NIV-INSACOG-GSEQ-588/2021** | EPI_ISL_1703962 | ICMR-National Institute of Virology - INSACOG | NIV Influenza | Dr. Varsha Potdar |
| **hCoV-19/Saudi Arabia/KFSHRC8/2021** | EPI_ISL_2151336 | Infectious Diseases, King Faisal Hospital Research Center | Infectious Diseases, King Faisal Hospital Research Cente | Alhamlan F,S., Al-Qahtani A,A., UdayaRaja G,K., Mutabagani M,S.,Balavenkatesh Mani,M., Althawadi S,I., Almaghrabi R,S., Alsanea M,S. and Alahideb B,M. |
| **hCoV-19/Hungary/HM-0122-362/2021** | EPI_ISL_1041203 | Hungarian Defence Forces Military Medical Centre | National Laboratory of Virology, Szentágothai Research Centre | Endre Gábor Tóth, Balázs Somogyi, Ágnes Balázs-Nagy, Csaba Pereszlényi,Ferenc Jakab, Gábor Kemenesi |
| **hCoV-19/Slovenia/90-017770-NM/2021** | EPI_ISL_1817500 | National Laboratory for Health, Environment and Food, OMM, Novo mesto | NLZOH (National Laboratory for Health, Environment and Food) / CISLD (Clinical Institute of Special Laboratory Diagnostics), University Children's Hospital, University | Sandra Janezic, Aleksander Mahnic, Maja Rupnik, Tjasa Žohar Čretnik, Alenka Štorman, Nika Gobec, Aleksander Kocuvan, Kaja Tominc, Maša Jarčič, David Cvetko, Tatjana Harlander, Matjaž Retelj / Jernej Kovač, Barbara Jenko Bizjan, Tine Tesovnik, |

| | | | | |
|---|---|---|---|---|
| | | | Medical Center Ljubljana | Robert Šket, Katarina Kozmos, Ana Grom, Maruša Debeljak, Marko Pokorn, Tadej Battelino |
| **hCoV-19/Armenia/IMB3-12/2021** | EPI_ISL_1718305 | National Center of Disease Control and Prevention of the Republic of Armenia | Institute of Molecular Biology NAS RA, Republic of Armenia, Department of Bioengineering, BioinformaticsInstitute and Molecular Biology IBMPh RAU, Republic of Armenia | Arsen Arakelyan, Diana Avetyan, Siras Hakobyan, Gisane Khachatyan, Maria Nikoghosyan, Tamara Sirunyan, Nelli Muradyan, Andranik Chavushyan, Hovsep Ghazaryan, Roksana Zakharyan, Shushan Sargrsyan, Gayane Melik-Pashayan |
| **hCoV-19/Jordan/AM-HU-13/2021** | EPI_ISL_1336799 | Prince Hamzah Hospital | Alanagreh | Loai Alanagreh, Mustafa Ababneh, Abdel-Ellah Al-Shudifat , Mai Ajluny, Hanan Abu alshaikh, Foad Alzoughool, Mohammad-Borhan Al-zghoul, Manar Atoum |
| **hCoV-19/Jordan/AM-HU-6/2021** | EPI_ISL_1336798 | Prince Hamzah Hospital | Alanagreh | Loai Alanagreh, Mustafa Ababneh, Abdel-Ellah Al-Shudifat , Mai Ajluny, Hanan Abu alshaikh, Foad Alzoughool, Mohammad-Borhan Al-zghoul, Manar Atoum |
| **hCoV-19/Denmark/DCGC-46965/2021** | EPI_ISL_1066476 | Department of Virus and Microbiological Special Diagnostics, Statens Serum Institut, Copenhagen, Denmark | Aalborg University | Danish Covid-19 Genome Consortium |
| **hCoV-19/Germany/BY-RKI-I-050934/2021** | EPI_ISL_1433396 | Labor Becker & Kollegen (Standort München) | IMS-10182-CVDP-D16F47A3-44F0-47F4-9443-E9A6C8E7D099 | Drechsel, Oliver |
| **hCoV-19/India/AP-CCMB-BJ395/2021** | EPI_ISL_1838367 | CSIR-Centre for Cellular and Molecular Biology | CSIR-Centre for Cellular and Molecular Biology-INSACOG | Payel Mukherjee,Lamuk Zaveri,Tulasi Nagabandi, Ara Sreenivas,Shreekant Verma, Amareshwar Vodapalli ,Blessy B John,Viswagithe S L,B Himasri,Valli Nagalakshmi Undamatla,Onkar Kulkarni,Sofia Banu,Archana Bharadwaj Siva,Sharath Chandra Thota,Karthik Bharadwaj Tallapaka,Rakesh K Mishra,Divya Tej Sowpati |

| | | | | |
|---|---|---|---|---|
| **hCoV-19/Germany/RP-RKI-I-056925/2021** | EPI_ISL_1440628 | SYNLAB MVZ Neustadt a. d. Weinstraße | Robert Koch Institute | Drechsel, Oliver |
| **hCoV-19/Wales/PHWC-PYYW9U/2021** | EPI_ISL_1476527 | Originating lab: Wales Specialist Virology Centre Sequencing lab: Pathogen Genomics Unit | Public Health Wales Microbiology Cardiff Wales Specialist Virology Centre | Catherine Moore, Johnathan Evans, Laura Gifford, Malorie Perry, Simon Cottrell, Angela Marchbank, Alec Birchley, Alexander Adams, Amy Gaskin, Bree Gatica-Wilcox, Jason Coombes, Joel Southgate, Lauren Gilbert, Lee Graham, Nicole Pacchiarini, Sara Kumziene-Summerhayes, Sarah Taylor, Sophie Jones, Sara Rey, Matthew Bull, Joanne Watkins, Sally Corden, Tom Connor |
| **hCoV-19/England/QEUH-12FDA6D/2021** | EPI_ISL_1256517 | Lighthouse Lab in Glasgow | Wellcome Sanger Institute for the COVID-19 Genomics UK (COG-UK) Consortium | Harper VanSteenhouse, Yumi Kasai, David Gray, Carol Clugston, Anna Dominiczak and Alex Alderton, Roberto Amato, Jeffrey Barrett, Sonia Goncalves, Ewan Harrison, David K. Jackson, Ian Johnston, Dominic Kwiatkowski, Cordelia Langford, John Sillitoe on behalf of the Wellcome Sanger Institute COVID-19 Surveillance Team |
| **hCoV-19/England/OXON-F481CE/2020** | EPI_ISL_951006 | Oxford Viromics, NDM, University of Oxford; Oxford University Hospitals; Basingstoke and North Hampshire Hospital | COVID-19 Genomics UK (COG-UK) Consortium | anya Golubchik, David Bonsall, George Macintyre, Amy Trebes, Mariateresa de Cesare, Catrin Moore, Alex Mobbs, Anita Justice, Robert Shaw, Monique Andersson, Timothy Peto, Emma Wise, Nathan Moore, Jessica Lynch, Nick Cortes, Matilde Mori, Stephen Kidd, David Buck, John Todd, Christophe Fraser |
| **hCoV-19/Scotland/OXON-F403B1/2020** | EPI_ISL_1833046 | Oxford Viromics, NDM, University of Oxford; Oxford University Hospitals; Basingstoke and North Hampshire Hospital | COVID-19 Genomics UK (COG-UK) Consortium | Tanya Golubchik, David Bonsall, George Macintyre, Amy Trebes, Mariateresa de Cesare, Catrin Moore, Alex Mobbs, Anita Justice, Robert Shaw, Monique Andersson, Timothy Peto, Emma Wise, Nathan Moore, Jessica Lynch, Nick Cortes, Matilde Mori, Stephen Kidd, David Buck, John Todd, |

| | | | | |
|---|---|---|---|---|
| | | | | Christophe Fraser |
| **hCoV-19/Spain/MD-59669058/2021** | EPI_ISL_2179789 | Servicio Microbiología Hospital La Paz | Servicio Microbiología Hospital La Paz | Fernando Lázaro, Rubén Cáceres, Jesús Mingorance Cruz, Elie Dahdouh |
| **hCoV-19/Hong Kong/HKPU-06804/2020** | EPI_ISL_1159106 | Department of Health Technology and Informatics, The Hong Kong Polytechnic University | Department of Health Technology and Informatics, The Hong Kong Polytechnic University | Gilman Kit-Hang Siu, Lam-Kwong Lee, Kenneth Siu-Sing Leung, Jake Siu-Lun Leung, Timothy Ting-Leung Ng, Chloe Toi-Mei Chan, Kingsley King-Gee Tam, Hiu-Yin Lao, Denise Sze-Hang Wong, Alan Ka-Lun Wu, Miranda Chong-Yee Yau, Yvette Wai-Man Lai, Kitty Sau-Chun Fung, Sandy Ka-Yee Chau, Barry Kin-Chung Wong, Wing-Kin To, Kristine Luk, Alex Yat-Man Ho, Tak-Lun Que, Kam-Tong Yip, Wing Cheong Yam, David Ho-Keung Shum, Shea Ping Yip |
| **hCoV-19/Finland/3228/2021** | EPI_ISL_2031006 | Department of Virology and Immunology, University of Helsinki and Helsinki University Hospital, Huslab Finland | Department of Virology, Faculty of Medicine, University of Helsinki, Helsinki, Finland | Teemu Smura, Ravi Kant, Phuoc Truong, Hussein Alburkat, Hannimari Kallio-Kokko, Jenni Virtanen, Maija Suvanto, Essi Korhonen, Sari Hannula, Harri Kangas, Hanna Liimatainen, Satu Kurkela, Hanna Jarva, Maija Lappalainen, Pekka Ellonen, Olli Vapalaht |
| **hCoV-19/Estonia/Cov11254/2021** | EPI_ISL_2642071 | SYNLAB Eesti OÜ | Department of Microbiology, Institute of Biomedicine and Translational Medicine, University of Tartu | Radko Avi, Aare Abroi, Irja Lutsar, Kristi Huik, Taavi Päll, Meri Pauskar, Ene-Ly Jõgeda, Arina Shablinskaja, Eveli Kallas, Kai Truusalu, Dagmar Hoidmets, Katrin Kaarna, Tuuli Reisberg, Lili Azin Milani, Ulvi Gerst Talas, Heiki Niglas, Olga Sadikova, Liidia Dotsenko, Mari-Anne Härma, Kaisa Truus, Mats Hansen, Paul Naaber, Andrio Lahesaare |
| **hCoV-19/Ireland/SO-NVRL-83IRL56730/2021** | EPI_ISL_909912 | National Virus Reference Laboratory | National Virus Reference Laboratory | Michael Carr, Gabriel Gonzalez, Jonathan Dean, Cillian F De Gascun |
| **hCoV-** | EPI_ISL_1272054 | Croatian Institute of Public Health | Croatian Institute of Public Health | Irena Tabain, Ivana Ferenčak |

| | | | | |
|---|---|---|---|---|
| **19/Croatia/561/2021** | | | | |
| **hCoV-19/Israel/CVL-2393/2021** | EPI_ISL_944546 | Israel Central Virology laboratory | Israel National Consortium for SARS-CoV-2 sequencing | Neta Zuckerman, Efrat Dahan Bucris, Michal Mandelboim, Dana Bar-Ilan, Oran Erster, Tzvia Mann, Omer Murik, David A. Zeevi, Assaf Rokney, Joseph Jaffe, Eva Nachum, Maya Davidovich Cohen, Ephraim Fass, Gal Zizelski Valenci, Mor Rubinstein, Efrat Rorman, Israel Nissan, Efrat Glick-Saar, Omri Nayshool, Gideon Rechavi, Ella Mendelson, Orna Mor |
| **hCoV-19/Finland/2737/2021** | EPI_ISL_1841818 | Department of Virology and Immunology, University of Helsinki and Helsinki University Hospital, Huslab Finland | Department of Virology, Faculty of Medicine, University of Helsinki, Helsinki, Finland | Teemu Smura, Ravi Kant, Phuoc Truong, Hussein Alburkat, Hannimari Kallio-Kokko, Jenni Virtanen, Maija Suvanto, Essi Korhonen, Sari Hannula, Harri Kangas, Hanna Liimatainen, Satu Kurkela, Hanna Jarva, Maija Lappalainen, Pekka Ellonen, Olli Vapalahti |
| **hCoV-19/Finland/V1070/2021** | EPI_ISL_1842531 | Department of Virology and Immunology, University of Helsinki and Helsinki University Hospital, Huslab Finland | Department of Virology, Faculty of Medicine, University of Helsinki, Helsinki, Finland | Teemu Smura, Ravi Kant, Phuoc Truong, Hussein Alburkat, Hannimari Kallio-Kokko, Jenni Virtanen, Maija Suvanto, Essi Korhonen, Sari Hannula, Harri Kangas, Hanna Liimatainen, Satu Kurkela, Hanna Jarva, Maija Lappalainen, Pekka Ellonen, Olli Vapalahti |
| **hCoV-19/Bahrain/02/2020** | EPI_ISL_486887 | National Influenza Center, Bahrain | National Influenza Center, Bahrain | Zaed,A., Altaif,Z., Shehab,F., AlWasti,H. |
| **hCoV-19/New Zealand/21MV0094/2021** | EPI_ISL_1082268 | LabPLUS | Institute of Environmental Science and Research (ESR) | Rachel Boyle, SallyAnn Harbison, Olivia Stroeven, Xiaoyun Ren, Matt Storey, Nikki Freed, Muhammad Faisal, Jing Wang, Hermes Perez, Anja Werno, Antje van der Linden, Arlo Upton, Chris Mansell, David Hammer, Dragana Drinkovic, Gary McAuliffe, Hana Sofia Andersson, James Ussher, Jill Sherwood, Josh Freeman, Julia |

| | | | | |
|---|---|---|---|---|
| | | | | Howard, Juliet Elvy, Mary DeAlmeida, Matt Blakiston, Matthew Rogers, Max Bloomfield, Michael Addidle, Michelle Balm, Sally Roberts, Sarah Jefferies, Sharmini Muttaiyah, Susan Morpeth, Susan Taylor, Timothy Blackmore, Vani Sathyendran, Veronica Playle, Virginia Hope, Erasmus Smit, Lauren Jelly, Olin Silander, Joep de Ligt |
| **hCoV-19/Austria/CeMM0018/2020** | EPI_ISL_419671 | Center for Virology, Medical University of Vienna | Bergthaler laboratory, CeMM Research Center for Molecular Medicine of the Austrian Academy of Sciences | Alexandra Popa, Benedikt Agerer, Henrique Colaco, Lukas Endler, Jakob-Wendelin Genger, Alexander Lercher, Mark Smyth, Thomas Penz, Michael Schuster, Judith Aberle, Stephan Aberle, Elisabeth Puchhammer-Stöckl, Christoph Bock, Andreas Bergthaler |
| **hCoV-19/Peru/AMA-UPCH-0261/2020** | EPI_ISL_729907 | Instituto de Medicina Tropical, Universidad Nacional Toribio Rodríguez de Mendoza de Amazonas | Laboratorio de Genómica Microbiana, Universidad Peruana Cayetano Heredia | Pablo Tsukayama, Alejandra Dávila-Barclay, Luis González, Pedro E. Romero, Brenda Ayzanoa, Janet Huancachoque, Pool Marcos, Stella Chenet, Rafael Tapia, Cecilia Pajuelo, Carla Montenegro |
| **hCoV-19/Australia/NSW1427/2021** | EPI_ISL_803111 | Sydney South West Pathology Service (SSWPS) - Royal Prince Alfred Hospital - NSW Health Pathology | NSW Health Pathology - Institute of Clinical Pathology and Medical Research; Westmead Hospital; University of Sydney | CIDM-PH et al. |
| **hCoV-19/Egypt/NRC-6465/2020** | EPI_ISL_2232367 | Center of Scientific Excellence for Influenza Viruses (CSEIV), National Research Centre | Center of Scientific Excellence for Influenza Viruses (CSEIV), National Research Centre | Rabeh El-Shesheny, Ahmed E Kayed, Ahmed El-Taweel, Mokhtar Gomaa, Sara Mahmoud, Yassmin Moatasim, Omnia Kutkat, Mina Kamel, Noura M Abo Shama, Mohamed El Sayes, Mahmoud Shehata, Ahmed Mostafa, Ahmed Kandeil, Richard Webby, Ghazi Kayali, Mohamed Ahmed Ali |
| **hCoV-19/Germany/BW-RKI-I-018088/2021** | EPI_ISL_1148250 | Bioscientia MVZ Labor Karlsruhe GmbH | Robert Koch Institute | Drechsel, Oliver |

| | | | | |
|---|---|---|---|---|
| **hCoV-19/South Africa/KRISP-MDSH920916/2020** | EPI_ISL_736927 | MDS | KRISP, KZN Research Innovation and Sequencing Platform | Giandhari J, Pillay S, Lessells R, ChimukangaraB, Mdlalose K, York D, Khan S, Tegally H, Wilkinson E, de Oliveira T |
| **hCoV-19/Saudi Arabia/KFSHRC1/2021** | EPI_ISL_ 2001099 | BIOSCIENCE, King Faisal Hospital Research Center | BIOSCIENCE, King Faisal Hospital Research Center | Alhamlan F,S., Al-Qahtani A,A., Mutabagani M,S., Althawadi S,I.,Almaghrabi R,S., Alahideb B,M., Alsanea M,S., UdayaRaja G,K. and Balavenkatesh Mani,M. |
| **hCoV-19/Australia/VIC13/2020** | EPI_ISL_419733 | Victorian Infectious Diseases Reference Laboratory (VIDRL) | Victorian Infectious Diseases Reference Laboratory and Microbiological Diagnostic Unit Public Health Laboratory, Doherty Institute | Caly L., Seemann T., Sait, M., Schultz M., Druce J., Sherry, N |
| **hCoV-19/Jordan/PHBC4/2021** | EPI_ISL_935043 | Biolab Diagnostic Laboratories | Princess Haya Biotechnology Center/ Jordan University of Science & Technology | Saied Jaradat, Hazem Haddad, Areej Alquran, Maha Karam, Shereen Issa,Suha Hasan, Amid Abdelnour, Issa Abu-Dayyeh |
| **hCoV-19/Bahrain/BAH-12/2020** | EPI_ISL_483553 | Kingdom of Bahrein Ministry of Health | Erasmus Medical Center | Bas Oude Munnink, David Nieuwenhuijse, Reina Sikkema, Fatema, Ebrahim Shehad, Amjad Ghanem Mohamed, Hashmeya Al Wasti, Claudia Schapendonk, Irina Chestakova, Anne van der Linden, Theo Bestebroer, Stefan van Nieuwkoop, Mark Pronk, Pascal Lexmond, Richard Molenkamp, Marion Koopmans, on behalf of the Dutch national COVID-19 response team. |
| **hCoV-19/New Zealand/20VR2572/2020** | EPI_ISL_579303 | Middlemore Hospital | Institute of Environmental Science and Research (ESR) | Xiaoyun Ren, Matt Storey, Nikki Freed, Muhammad Faisal, Jing Wang, Hermes Perez, Anja Werno, Antje van der Linden, Arlo Upton, Chris Mansell, David Hammer, Dragana Drinkovic, Gary McAuliffe, Hana Sofia Andersson, James Ussher, Jill Sherwood, Josh Freeman, Julia Howard, Juliet Elvy, Mary DeAlmeida, Matt |

| | | | | |
|---|---|---|---|---|
| | | | | Blakiston, Matthew Rogers, Max Bloomfield, |
| **hCoV-19/Chile/ML-ISPCH-1/2020** | EPI_ISL_414577 | Hospital de Talca, Chile | Instituto de Salud Publica de Chile | Andrés E. Castillo, Bárbara Parra, Paz Tapia, Alejandra Acevedo, Jaime Lagos, Winston Andrade, Loredana Arata, Gabriel Leal, Gisselle Barra, Carolina Tambley, Javier Tognarelli, Patricia Bustos, Soledad Ulloa, Rodrigo Fasce, Jorge Fernández. |
| **hCoV-19/New Zealand/20VR3156/2020** | EPI_ISL_579446 | Canterbury Health Laboratories | Institute of Environmental Science and Research (ESR) | Xiaoyun Ren, Matt Storey, Nikki Freed, Muhammad Faisal, Jing Wang, Hermes Perez, Anja Werno, Antje van der Linden, Arlo Upton, Chris Mansell, David Hammer, Dragana Drinkovic, Gary McAuliffe, Hana Sofia Andersson, James Ussher, |
| **hCoV-19/Norway/1378/2020** | EPI_ISL_2549107 | Oslo University Hospital, Department of Medical Microbiology | Norwegian Institute of Public Health, Department of Virology | Kathrine Stene-Johansen, Kamilla Heddeland Instefjord, Hilde Elshaug, Garcia Llorente Ignacio, Jon Bråte, Engebretsen Serina Beate,Pedersen Benedikte Nevjen, Line Victoria Moen, Debech Nadia, Atiya R Ali,Marie Paulsen Madsen, Rasmus Riis Kopperud, Hilde Vollan, Karoline Bragstad, Olav Hungnes |
| **hCoV-19/Germany/BW-RKI-I-050580/2021** | EPI_ISL_1432905 | Eurofins LifeCodexx GmbH | Robert Koch Institute | Drechsel, Oliver |
| **hCoV-19/Germany/BY-RKI-I-074312/2021** | EPI_ISL_1640642 | Eurofins LifeCodexx GmbH | Robert Koch Institute | Drechsel, Oliver |
| **hCoV-19/Iraq BAS-2/2020** | EPI_ISL_907075 | Department of Biology, University of Basrah | Department of Biology, University of Basrah | Abu-Ali,H.M. and Al-Badran,I.F. |
| **hCoV-19/England/20099000504/2020** | EPI_ISL_464200 | Respiratory Virus Unit, Microbiology Services Colindale, Public Health England | Respiratory Virus Unit, Microbiology Services Colindale, Public Health England | PHE Covid Sequencing Team |

| | | | | |
|---|---|---|---|---|
| **hCoV-19/France/BFC-IPP00708/2021** | EPI_ISL_872282 | Labo Analyses Med | National Reference Center for Viruses of Respiratory Infections, Institut Pasteur, Paris | Marion Barbet, Sylvie Behillil, Méline Bizard, Angela Brisebarre, Camille Capel, Etienne Simon-Lorière, Vincent Enouf, Maud Vanpeene, Sylvie van der Werf,Girard Sophie |
| **hCoV-19/Mexico/CMX-INER-IMSS-00184/2021** | EPI_ISL_1279455 | Laboratorio Central de Epidemiología (LCE) | Instituto Nacional de Enfermedades Respiratorias (INER): Centro de Investigación en Enfermedades Infecciosas (CIENI) | Consorcio Mexicano de Vigilancia Genómica (CoViGen-Mex). Authors (in alphabetical order): Julio Elias Alvarado-Yaah, Carlos F. Arias, Santiago Ávila-Ríos, Víctor Hugo Borja-Aburto, Celia Boukadida, Juan Bautista Chale-Dzul , José Antonio Enciso-Moreno, Gloria Elena Espinoza-Ayala, Fernando Fontove-Herrera, Concepción Grajales-Muñiz, Ricardo Grande, Alfredo Herrera-Estrella, |
| **hCoV-19/England/CAMC-B218D9/2020** | EPI_ISL_647134 | Lighthouse Lab in Cambridge | Wellcome Sanger Institute for the COVID-19 Genomics UK (COG-UK) Consortium | Rob Howes, The Lighthouse Lab in Cambridge and Alex Alderton, Roberto Amato, Sonia Goncalves, Ewan Harrison, David K. Jackson, Ian Johnston, Dominic Kwiatkowski, Cordelia Langford, John Sillitoe on behalf of the Wellcome Sanger Institute COVID-19 Surveillance Team |
| **hCoV-19/Saudi Arabia/KAUST-Madinah269/2020** | EPI_ISL_437751 | Pathogen Genomics Lab King Abdullah University of Science and Technology(KAUST) | Pathogen Genomics Lab King Abdullah University of Science and Technology(KAUST) | Sharif Hala,Fadwa Alofi,Afrah Alsomali, Asim Khogeer, Sara Mfarrej, Khaled Algithami,Raeece Naeem, Amit Kumar Subudhi,Fathia Ben-Rached, Rahul Salunke, Anwar Hashem, Naif Almontashiri, Arnab Pain |
| **hCoV-19/Turkey/HSGM-10992/2021** | EPI_ISL_1790199 | Ministry of Health Turkey | Ministry of Health Turkey | Fatma Bayrakdar, Yasemin Cosgun, Suleyman Yalcin, Gulay Korukluoglu |
| **hCoV-19/USA/PA-CDC-STM-000052590/2021** | EPI_ISL_1796860 | Helix/Illumina | Centers for Disease Control and Prevention Division of Viral Diseases, Pathogen Discovery | Dakota Howard, Dhwani Batra, Peter W. Cook, Kara Moser, Adrian Paskey, Jason Caravas, Benjamin Rambo-Martin, Shatavia Morrison, Christopher Gulvick, Scott Sammons, Yvette Unoarumhi, Darlene |

| | | | | |
|---|---|---|---|---|
| | | | | Wagner, Matthew Schmerer, Eileen de Feo, Jan Antico, |
| **hCoV-19/Israel/CVL-1461/2021** | EPI_ISL_1210225 | Israel Central Virology laboratory | Israel National Consortium for SARS-CoV-2 sequencing | Neta Zuckerman, Efrat Dahan Bucris, Michal Mandelboim, Dana Bar-Ilan, Oran Erster, Tzvia Mann, Omer Murik, David A. Zeevi, Assaf Rokney, Joseph Jaffe, Eva Nachum, Maya Davidovich Cohen, Ephraim Fass, Gal Zizelski Valenci, Mor Rubinstein, Efrat Rorman, Israel Nissan, Efrat Glick-Saar, Omri Nayshool, Gideon Rechavi, Ella Mendelson, Orna Mor |
| **hCoV-19/USA/TX-HMH-MCoV-35772/2021** | EPI_ISL_2201206 | Houston Methodist Hospital | Houston Methodist Hospital | Randall J. Olsen, Paul A. Christensen, S. Wesley Long, Sishir Subedi, Robert Olson, Marcus Nguyen, James J. Davis, Matthew Ojeda Saavedra, Prasanti Yerramilli, Layne Pruitt, Kristina Reppond, Madison N. Shyer, Jessica Cambric, Ryan Gadd, Ilya J. Finkelstein, Jimmy Gollihar, and James M. Musser |
| **hCoV-19/Aruba/AW-RIVM-10757/2021** | EPI_ISL_1059915 | Dutch COVID-19 response team | National Institute for Public Health and the Environment (RIVM) | Adam Meijer, Harry Vennema, Dirk Eggink, Jeroen Cremer, Sharon van den Brink, Bas van der Veer, AnneMarie van den Brandt, Florian Zwagemaker, Dennis Schmitz, Chantal Reusken, on behalf of the national COVID-19 response team |
| **hCoV-19/Australia/QLD1470/2020** | EPI_ISL_1300530 | Public Health Virology-Forensic and Scientific Services (PHV-FSS) | Public Health Virology-Forensic and Scientific Services (PHV-FSS) | Son Nguyen |
| **hCoV-19/France/CVL-IPP00317/2021** | EPI_ISL_860909 | CHU Tours - Virologie | National Reference Center for Viruses of Respiratory Infections, Institut Pasteur, Paris | Marion Barbet, Sylvie Behillil, Méline Bizard, Angela Brisebarre, Camille Capel, Etienne Simon-Lorière, Vincent Enouf, Maud Vanpeene, Sylvie van der Werf,Gaudy Graffin Catherine |
| **hCoV-19/Turkey/HSGM-** | EPI_ISL_2157882 | Ministry of Health Turkey | Ministry of Health Turkey | Fatma Bayrakdar, Yasemin Cosgun, Suleyman Yalcin, Gulay Korukluoglu |

| | | | | |
|---|---|---|---|---|
| **11079/2021** | | | | |
| **hCoV-19/Iraq/PCRLab-01/2021** | EPI_ISL_2379893 | The teaching Baghdad hospital | PCR lab | Abdulhussein Thair, Fadhil Hula |
| **hCoV-19/Austria/CeMM1083/2020** | EPI_ISL_583885 | Austrian Agency for Health and Food Safety (AGES | Bergthaler laboratory, CeMM Research Center for Molecular Medicine of the Austrian Academy of Sciences | Alexandra Popa, Benedikt Agerer, Henrique Colaco, Lukas Endler, Jakob-Wendelin Genger, Alexander Lercher, Mark Smyth, Thomas Penz, Michael Schuster, Jan Laine, Martin Senekowitsch, Judith Aberle, Stephan Aberle, Peter Hufnagl, Daniela Schmid |
| **hCoV-19/Ireland/WD-NVRL-71IRL06472/2020** | EPI_ISL_500582 | National Virus Reference Laboratory | National Virus Reference Laboratory | Michael Carr, Gabriel Gonzalez, Jonathan Dean, Suzie Coughlan, Cillian F De Gascun |
| **hCoV-19/Norway/3584/2020** | EPI_ISL_635186 | University Hospital of Northern Norway, Department for Microbiology and Infectious Disease Control | Norwegian Institute of Public Health, Department of Virology | Kathrine Stene-Johansen, Kamilla Heddeland Instefjord, Hilde Elshaug, Marie Paulsen Madsen, Rasmus Riis Kopperud, Hilde Vollan, Karoline Bragstad, Olav Hungnes |
| **hCoV-19/Afghanistan/IMB07962/2020** | EPI_ISL_1001000 | Bundeswehr Institute of Microbiology | Bundeswehr Institute of Microbiology | Markus Antwerpen, Alexandra Rehn, Mathias Walter, Malena Bestehorn-Willmann, Sabine Zange, Enrico Georgi, Roman Wölfel |
| **hCoV-19/Portugal/PT5110/2021** | EPI_ISL_1494779 | Centro Hospitalar de Entre o Douro e Vouga (CHEDV) | Institute of Biomedicine (IBiMED), Universidade de Aveiro | Gabriela Moura, Sofia Marques, Patricia Arinto, Miguel Pinheiro and Manuel Santos |
| **hCoV-19/Denmark/DCGC-17661/2020** | EPI_ISL_751130 | Department of Virus and Microbiological Special Diagnostics, Statens Serum Institut, Copenhagen, Denmark | Albertsen Lab, Department of Chemistry and Bioscience, Aalborg University, Denmark | Danish Covid-19 Genome Consortium |
| **hCoV-19/England/ALDP-** | EPI_ISL_675632 | Lighthouse Lab in Alderley Park | Wellcome Sanger Institute for the COVID-19 Genomics UK (COG-UK) | Jacquelyn Wynn, Mairead Hyland, The Lighthouse Lab in Alderley Park and Alex |

| | | | | |
|---|---|---|---|---|
| **B83161/2020** | | | Consortium | Alderton, Roberto Amato, Sonia Goncalves, Ewan Harrison, David K. Jackson, Ian Johnston, Dominic Kwiatkowski, Cordelia Langford, John Sillitoe on behalf of the Wellcome Sanger Institute COVID-19 Surveillance Team |
| **hCoV-19/Iraq/ICGEB-5T/2020** | EPI_ISL_582030 | Biology Department, College of Science, Al-Muthanna University | International Centre for Genetic Engineering and Biotechnology (ICGEB) and ARGO Open Lab Platform | Nihad Al-Rashedi, Danilo Licastro, Sreejith Rajasekharan, Simeone Dal Monego, Alessandro Marcello |
| **hCoV-19/Belgium/UZA-UA-CV2008146136/2021** | EPI_ISL_1190853 | Platform BIS UZA/UAntwerpen | UAntwerp, Laboratory of Medical Microbiology | Basil Britto Xavier, Jasmine Coppens, Marie Le Mercier, Christine Lammens, Veerle Matheeussen, Herman Goossens |
| **hCoV-19/Jordan/Biolab020/2021** | EPI_ISL_1823202 | Biolab Diagnostic Laboratories | Biolab Diagnostic Laboratories | Issa Abu-Dayyeh, Ahmad Tibi, Lama Hussein, Shayma Ali, Badia Saddedin, Eiad Atwa, Amid Abdelnour |
| **hCoV-19/USA/GA-GPHL-0057/2021** | EPI_ISL_1262463 | GA Department of Public Health | GA Department of Public Health | Stacy Reeves, Jonathan Edwards, Cynthia Dixey, Tonia Parrott, Aliyah Fields, Taylor Smith |
| **hCoV-19/New Zealand/20CV0669/2020** | EPI_ISL_732969 | Middlemore Hospital | Institute of Environmental Science and Research (ESR) | Xiaoyun Ren, Matt Storey, Nikki Freed, Muhammad Faisal, Jing Wang, Hermes Perez, Anja Werno, Antje van der Linden, Arlo Upton, Chris Mansell, David Hammer, Dragana Drinkovic, Gary McAuliffe, Hana Sofia Andersson, James Ussher, Jill Sherwood, Josh Freeman, Julia Howard, Juliet Elvy, Mary DeAlmeida, Matt Blakiston, Matthew Rogers, Max Bloomfield, Michael Addidle, Michelle Balm, Sally Roberts, Sarah Jefferies, Sharmini Muttaiyah, Susan Morpeth, Susan Taylor, Timothy Blackmore, Vani Sathyendran, Veronica Playle, Virginia |

| | | | | |
|---|---|---|---|---|
| | | | | Hope, Erasmus Smit, Lauren Jelly, Olin Silander, Joep de Ligt |
| **hCoV-19/Kuwait/SKS00103-JAH/2020** | EPI_ISL_903389 | MOH - Jaber Al-Ahmad Hospital (Innovation Research Laboratory) | MOH - Jaber Al-Ahmad Hospital (Innovation Research Laboratory) | Salman Al-Sabah , Mohammad Alghounaim |
| **hCoV-19/Iraq ICGEB-2T/2020** | EPI_ISL_582029 | Biology Department, College of Science, Al-Muthanna University | International Centre for Genetic Engineering and Biotechnology (ICGEB) and ARGO Open Lab Platform | Nihad Al-Rashedi, Danilo Licastro, Sreejith Rajasekharan, Simeone Dal Monego, Alessandro Marcello |
| **hCoV-19/South Africa/VIDA-KRISP-V001019/2020** | EPI_ISL_940877 | Vaccines and Infectious Diseases Analytics Research Unit (VIDA | KRISP, KZN Research Innovation and Sequencing Platform | Baillie Vicky, du Plessis Jeanine, Giandhari Jennifer, Pillay Sureshnee, Naidoo Yeshnee, Tegally Houriiyah, de Oliveira Tulio, Madhi Shabir |
| **hCoV-19/USA/MA-MASPHL-03369/2021** | EPI_ISL_2008374 | Massachusetts State Public Health Laboratory | Massachusetts State Public Health Laboratory | Andrew Lang, Timelia Fink, Glen Gallagher, Sandra Smole |
| **hCoV-19/Finland/4992/2021** | EPI_ISL_2259002 | Department of Virology and Immunology, University of Helsinki and Helsinki University Hospital, Huslab Finland | Department of Virology, Faculty of Medicine, University of Helsinki, Helsinki, Finland | Teemu Smura, Ravi Kant, Phuoc Truong, Hussein Alburkat, Hannimari Kallio-Kokko, Jenni Virtanen, Maija Suvanto, Essi Korhonen, Sari Hannula, Harri Kangas, Hanna Liimatainen, Satu Kurkela, Hanna Jarva, Maija Lappalainen, Pekka Ellonen, Olli Vapalahti |
| **hCoV-19/France/HDF-P037-21144M1410/2021** | EPI_ISL_1818807 | CH Lens | CHU Lille - Laboratoire de Virologie | AIT YAHYA Emilie, ALIDJINOU Enagnon Kazali, BOCKET Laurence, CREPIN Michel, DEMAY Christophe, ENGELMANN Ilka, GEFFROY Sandrine, GUIGON Aurélie, LAMBERT Valérie, LAZREK Mouna, NOBILLIAUX Florian, PREVOST Brigitte, THUILLIER Caroline, TINEZ Claire |
| **hCoV-19/South Africa/KRISP-EG00467095/2020** | EPI_ISL_736954 | NHLS-IALCH | KRISP, KZN Research Innovation and Sequencing Platform | Giandhari J, Pillay S, Lessells R, ChimukangaraB, Mdlalose K, York D, Khan S, Tegally H, Wilkinson E, de Oliveira T |

| | | | | |
|---|---|---|---|---|
| **hCoV-19/Saudi Arabia/KFSHRC10/2021** | EPI_ISL_2151337 | Infectious Diseases, King Faisal Hospital Research Center | Infectious Diseases, King Faisal Hospital Research Center | Alhamlan F,S., Al-Qahtani A,A., Mutabagani M,S., Althawadi S,I.,Almaghrabi R,S., Alahideb B,M., Alsanea M,S., Balavenkatesh Mani,M. and UdayaRaja G,K. |
| **hCoV-19/Iraq/BAS-1/2020** | EPI_ISL_956332 | Department of Biology, University of Basrah | Department of Biology, University of Basrah | Abu-Ali,H.F. and Al-Badran,A.I. |
| **hCoV-19/Iraq/USAFSAM-S092/2020** | EPI_ISL_812285 | Landstuhl Regional Medical Center | United States Air Force School of Aerospace Medicine | Anthony Fries, Jennifer Meyer, Amanda Javorina, Sarah Purves, William Gruner, Clarise Starr, Elizabeth Macias, Fritz Castillo, Cole Anderson |
| **hCoV-19/Jordan/SR-039/2020** | EPI_ISL_429998 | Biolab Diagnostic Laboratories | Andersen lab at Scripps Research | Issa Abu-Dayyeh, Ahmad Tibi, Lama Hussein, Lina Mohammad, Zein Naber, Amid Abdelnour with SEARCH Alliance San Diego |
| **hCoV-19/New Zealand/20CV0291/2020** | EPI_ISL_579081 | Canterbury Health Laboratories | Institute of Environmental Science and Research (ESR) | Xiaoyun Ren, Matt Storey, Nikki Freed, Muhammad Faisal, Jing Wang, Hermes Perez, Anja Werno, Antje van der Linden, Arlo Upton, Chris Mansell, David Hammer, Dragana Drinkovic, Gary McAuliffe, Hana Sofia Andersson, James Ussher, Jill Sherwood, Josh Freeman, Julia Howard, Juliet Elvy, Mary DeAlmeida, Matt Blakiston, Matthew Rogers, Max Bloomfield, Michael Addidle, Michelle Balm, Sally Roberts, Sarah Jefferies, Sharmini Muttaiyah, Susan Morpeth, Susan Taylor, Timothy Blackmore, Vani Sathyendran, Veronica Playle, Virginia Hope, Erasmus Smit, Lauren Jelly, Olin Silander, Joep de Ligt |
| **hCoV-19/Saudi Arabia/KAUST-MADINAH1655/2020** | EPI_ISL_751235 | Pathogen Genomics Lab King Abdullah University of Science and Technology(KAUST) | Pathogen Genomics Lab King Abdullah University of Science and Technology(KAUST) | Sara Mfarrej, Olga Douvropoulou, Raushan Nugmanova, Raeece Naeem, Sharif Hala, Fadwa Alofi, Asim Khogeer, Afrah Alsomali, Jumana Taha, Abdulaziz Alahmadi, Kahled Algithami, Anwar Hashem, Naif Almontashiri, Arnab Pain |

| | | | | |
|---|---|---|---|---|
| **hCoV-19/Hong Kong/HKPU-00248/2020** | EPI_ISL_1289436 | Department of Health Technology and Informatics, The Hong Kong Polytechnic University | Department of Health Technology and Informatics, The Hong Kong Polytechnic University | Gilman Kit-Hang Siu, Lam-Kwong Lee, Kenneth Siu-Sing Leung, Jake Siu-Lun Leung, Timothy Ting-Leung Ng, Chloe Toi-Mei Chan, Kingsley King-Gee Tam, Hiu-Yin Lao, Denise Sze-Hang Wong, Alan Ka-Lun Wu, Miranda Chong-Yee Yau, Yvette Wai-Man Lai, Kitty Sau-Chun Fung, Sandy Ka-Yee Chau, Barry Kin-Chung Wong, Wing-Kin To, Kristine Luk, Alex Yat-Man Ho, Tak-Lun Que, Kam-Tong Yip, Wing Cheong Yam, David Ho-Keung Shum, Shea Ping Yip |
| **hCoV-19/Spain/CL-COV05707/2021** | EPI_ISL_2986426 | SARS-CoV-2 Sequencing Castilla y Leon-Spain Consortium | SARS-CoV-2 Sequencing Castilla y Leon-Spain Consortium | Antonio Orduña-Domingo, Marta Hernandez, David Abad, Marta Dominguez-Gil, Silvia Rojo, Gabriel March Rossello, Sonsoles Garcinuño Pérez, Carmen Aldea-Mansilla, Mª Fe Brezmes-Valdivieso, Gregoria Megías Lobón, María Antonia García Castro, Carmen Gimeno Crespo, Noelia Arenal Andrés, Carlos Fuster Foz, M. Isabel Fernandez-Natal, Jose María Eiros Bouza |
| **hCoV-19/USA/PA-VSP2225/2021** | EPI_ISL_2597536 | Hospital of the University of Pennsylvania Molecular Pathology Lab | Bushman Lab - University of Pennsylvania | John K. Everett, Kyle Rodino, Shantan Reddy, Aoife M. Roche, Young Hwang, Scott Sherrill-Mix, Samantha A. Whiteside, Jevon Graham-Wooten, Layla A. Khatib, Ayannah S. Fitzgerald, Arupa Ganguly, Mike Feldman, Brendan Kelly, Ronald G. Collman and Frederic Bushan |
| **hCoV-19/Iraq/Erbil-1/2021** | EPI_ISL_2153105 | GENETICS DEPARTMENT, ZHEEN INTERNATIONAL HOSPITAL | GENETICS DEPARTMENT, ZHEEN INTERNATIONAL HOSPITAL | Khailany,R.A., Rahman,M.O., Ozaslan,M. |
| **hCoV-19/Germany/BW-RKI-I-043354/2021** | EPI_ISL_1350542 | Limbach - MVZ Humangenetik Ulm | IMS-10146-CVDP-1204805D-6911-4D4C-86D5-02091960ED49 | Drechsel, Oliver |
| **hCoV-19/India/PB-ICMR-NIV-INSACOG-** | EPI_ISL_1703962 | ICMR-National Institute of Virology - | NIV Influenza | Dr. Varsha Potdar |

| | | | | |
|---|---|---|---|---|
| **GSEQ-588/2021** | | INSACOG | | |
| **hCoV-19/England/PHEC-159966/2021** | EPI_ISL_2737121 | Respiratory Virus Unit, Microbiology Services Colindale, Public Health England | COVID-19 Genomics UK (COG-UK) Consortium | PHE Covid Sequencing Team |
| **hCoV-19/USA/OH-UHTL-992/2021** | EPI_ISL_2661529 | UHTL, University Hospitals | UHTL, University Hospitals | Sadri,N., Alouani,D., Song,X. |
