## Supplementary material for "High prevalence of an alpha variant lineage with a premature stop codon in ORF7a in Iraq, winter 2020-2021": Table S2

### Supplementary materials

**Table S2**: A set of aa changes located in spike

| **Virus name** | **Spike aa changes** | **GISAID clade** |
|---|---|---|
| hCoV-19/Iraq/Samawa-4/2021<br>hCoV-19/Iraq/Samawa-7/2021<br>hCoV-19/Iraq/Samawa-14/2021<br>hCoV-19/Iraq/Samawa-22/2021 | H69del, V70del, Y144del, G181A, N501Y, A570D, D614G, P681H, T716I, S982A, D1118H | GRY |
| hCoV-19/Iraq/Samawa-1/2021<br>hCoV-19/Iraq/Samawa-2/2021<br>hCoV-19/Iraq/Samawa-3/2020<br>hCoV-19/Iraq/Samawa-5/2021<br>hCoV-19/Iraq/Samawa-6/2021<br>hCoV-19/Iraq/Samawa-8/2021<br>hCoV-19/Iraq/Samawa-9/2021<br>hCoV-19/Iraq/Samawa-10/2021<br>hCoV-19/Iraq/Samawa-12/2021<br>hCoV-19/Iraq/Samawa-13/2021<br>hCoV-19/Iraq/Samawa-15/2020<br>hCoV-19/Iraq/Samawa-16/2021<br>hCoV-19/Iraq/Samawa-19/2021<br>hCoV-19/Iraq/Samawa-20/2021<br>hCoV-19/Iraq/Samawa-21/2021<br>hCoV-19/Iraq/Samawa-23/2021<br>hCoV-19/Iraq/Samawa-24/2021<br>hCoV-19/Iraq/Samawa-27/2021<br>hCoV-19/Iraq/Samawa-28/2021<br>hCoV-19/Iraq/Samawa-32/2021<br>hCoV-19/Iraq/Samawa-33/2021<br>hCoV-19/Iraq/Samawa-34/2021<br>hCoV-19/Iraq/Samawa-35/2021<br>hCoV-19/Iraq/Samawa-39/2021<br>hCoV-19/Iraq/Samawa-41/2021<br>hCoV-19/Iraq/Samawa-42/2021<br>hCoV-19/Iraq/Samawa-48/2021<br>hCoV-19/Iraq/Samawa-51/2021 | H69del, V70del, Y144del, N501Y, A570D, D614G, P681H, T716I, S982A, D1118H | GRY |
| hCoV-19/Iraq/Samawa-53/2021 | Y144del, N501Y, A570D, D614G, P681H, T716I, S982A, D1118H | GR |
| hCoV-19/Iraq/Samawa-78/2021 | V70L, N501Y, A570D, D614G, P681H, T716I, S982A, D1118H | GR |
| hCoV-19/Iraq/Samawa-17/2021 | S151T, D614G, Q675H | GH |

| | | |
|---|---|---|
| hCoV-19/Iraq/Samawa-18/2021 | L452M, D614G | GH |
| hCoV-19/Iraq/Samawa-19/2021 | L5F, H69del, V70del, Y144del, N501Y, A570D, D614G, P681H, T716I, S982A, D1118H | GRY |
| hCoV-19/Iraq/Samawa-25/2021<br>hCoV-19/Iraq/Samawa-26/2021<br>hCoV-19/Iraq/Samawa-29/2021<br>hCoV-19/Iraq/Samawa-30/2021<br>hCoV-19/Iraq/Samawa-31/2021<br>hCoV-19/Iraq/Samawa-40/2021<br>hCoV-19/Iraq/Samawa-45/2021<br>hCoV-19/Iraq/Samawa-46/2021<br>hCoV-19/Iraq/Samawa-47/2021<br>hCoV-19/Iraq/Samawa-56/2021<br>hCoV-19/Iraq/Samawa-58/2021<br>hCoV-19/Iraq/Samawa-60/2021<br>hCoV-19/Iraq/Samawa-62/2021<br>hCoV-19/Iraq/Samawa-63/2021<br>hCoV-19/Iraq/Samawa-64/2021<br>hCoV-19/Iraq/Samawa-65/2021<br>hCoV-19/Iraq/Samawa-67/2021 | H69del, V70del, Y144del , A570D, D614G, P681H, T716I, S982A, D1118H | GR |
| hCoV-19/Iraq/Samawa-73/2021 | A570D, D614G, S982A, D1118H | GR |
| hCoV-19/Iraq/Samawa-72/2021 | A570D, D614G, P681H, D1118H | GR |
| hCoV-19/Iraq/Samawa-55/2021<br>hCoV-19/Iraq/Samawa-59/2021<br>hCoV-19/Iraq/Samawa-61/2021<br>hCoV-19/Iraq/Samawa-66/2021 | A570D, D614G, S982A | GR |
| hCoV-19/Iraq/Samawa-36/2021 | D614G, P681H, S982A | GR |
| hCoV-19/Iraq/Samawa-37/2021<br>hCoV-19/Iraq/Samawa-38/2021 | D614G, Q675H | GH |
| hCoV-19/Iraq/Samawa-43/2021 | D614G, S982A | GR |
| hCoV-19/Iraq/Samawa-44/2021 | D614G, P681H, T716I, S982A, D1118H | GR |
| hCoV-19/Iraq/Samawa-49/2021 | L18F, A222V | O |
| hCoV-19/Iraq/Samawa-50/2021 | T19R, L452Q, D614G | GR |
| hCoV-19/Iraq/Samawa-52/2021 | D80A D215G L242del A243del L244del E484K N501Y D614G A701V | GH |
