## Supplementary material for "High prevalence of an alpha variant lineage with a premature stop codon in ORF7a in Iraq, winter 2020-2021": Table S3

### Supplementary materials

**Table** S3:

| Isolate name | Infection Severity | Patient Status | ORF7a Stop | GenBank ID |
|---|---|---|---|---|
| hCoV-19/Iraq/Samawa-1/2021 | Moderate | Deceased | - | MZ145303 |
| hCoV-19/Iraq/Samawa-2/2021 | Moderate | Live | 27577* | MZ145309 |
| hCoV-19/Iraq/Samawa-3/2020 | Moderate | Live | 27577* | MZ145318 |
| hCoV-19/Iraq/Samawa-4/2021 | Moderate | Live | 27577* | MZ145324 |
| hCoV-19/Iraq/Samawa-5/2021 | Moderate | Live | 27577* | MZ145327 |
| hCoV-19/Iraq/Samawa-6/2021 | Mild | Live | 27577* | MZ145329 |
| hCoV-19/Iraq/Samawa-7/2021 | Mild | Live | 27577* | MZ145330 |
| hCoV-19/Iraq/Samawa-8/2021 | Severe | Live | 27577* | MZ145334 |
| hCoV-19/Iraq/Samawa-9/2021 | Mild | Live | 27577* | MZ145338 |
| hCoV-19/Iraq/Samawa-10/2021 | Moderate | Live | 27577* | MZ145296 |
| hCoV-19/Iraq/Samawa-11/2021 | Moderate | Live | 27577* | MZ145297 |
| hCoV-19/Iraq/Samawa-12/2021 | Mild | Live | 27577* | MZ145298 |
| hCoV-19/Iraq/Samawa-13/2021 | Moderate | Live | - | MZ145299 |
| hCoV-19/Iraq/Samawa-14/2021 | Severe | Deceased | 27577* | MZ145300 |
| hCoV-19/Iraq/Samawa-15/2020 | Mild | Live | - | MZ145301 |
| hCoV-19/Iraq/Samawa-16/2021 | Mild | Live | 27577* | MZ145302 |
| hCoV-19/Iraq/Samawa-17/2021 | Severe | Live | - | MZ145291 |
| hCoV-19/Iraq/Samawa-18/2021 | Moderate | Live | - | MZ145288 |
| hCoV-19/Iraq/Samawa-19/2021 | Mild | Live | 27577* | MZ145304 |
| hCoV-19/Iraq/Samawa-20/2021 | Mild | Live | 27577* | MZ145305 |
| hCoV-19/Iraq/Samawa-21/2021 | Moderate | Live | - | MZ145306 |
| hCoV-19/Iraq/Samawa-22/2021 | Mild | Live | 27577* | MZ145307 |
| hCoV-19/Iraq/Samawa-23/2021 | Mild | Live | 27577* | MZ145308 |
| hCoV-19/Iraq/Samawa-24/2021 | Severe | Live | - | MZ145310 |

| | | | | |
|---|---|---|---|---|
| hCoV-19/Iraq/Samawa-25/2021 | Moderate | Live | 27577* | MZ145311 |
| hCoV-19/Iraq/Samawa-26/2021 | Moderate | Live | 27577* | MZ145312 |
| hCoV-19/Iraq/Samawa-27/2021 | Mild | Live | 27577* | MZ145313 |
| hCoV-19/Iraq/Samawa-28/2021 | Mild | Live | 27577* | MZ145314 |
| hCoV-19/Iraq/Samawa-29/2021 | Mild | Live | - | MZ145315 |
| hCoV-19/Iraq/Samawa-30/2021 | Moderate | Live | - | MZ145316 |
| hCoV-19/Iraq/Samawa-31/2021 | Moderate | Live | - | MZ145317 |
| hCoV-19/Iraq/Samawa-32/2021 | Moderate | Live | - | MZ145319 |
| hCoV-19/Iraq/Samawa-33/2021 | Moderate | Live | - | MZ145320 |
| hCoV-19/Iraq/Samawa-34/2021 | Moderate | Live | - | MZ145321 |
| hCoV-19/Iraq/Samawa-35/2021 | Moderate | Live | - | MZ145322 |
| hCoV-19/Iraq/Samawa-36/2021 | Mild | Live | 27577* | MZ145289 |
| hCoV-19/Iraq/Samawa-37/2021 | Mild | Live | - | MZ145290 |
| hCoV-19/Iraq/Samawa-38/2021 | Moderate | Live | - | MZ145294 |
| hCoV-19/Iraq/Samawa-39/2021 | Mild | Live | 27577* | MZ145323 |
| hCoV-19/Iraq/Samawa-40/2021 | Moderate | Live | - | MZ145325 |
| hCoV-19/Iraq/Samawa-41/2021 | Moderate | Live | - | MZ145326 |
| hCoV-19/Iraq/Samawa-42/2021 | Mild | Live | - | MZ145328 |
| hCoV-19/Iraq/Samawa-43/2021 | Mild | Live | - | MZ145287 |
| hCoV-19/Iraq/Samawa-44/2021 | Moderate | Live | - | MZ145331 |
| hCoV-19/Iraq/Samawa-45/2021 | Moderate | Live | 27577* | MZ145332 |
| hCoV-19/Iraq/Samawa-46/2021 | Mild | Live | - | MZ145333 |
| hCoV-19/Iraq/Samawa-47/2021 | Mild | Live | - | MZ145335 |
| hCoV-19/Iraq/Samawa-48/2021 | Moderate | Live | 27577* | MZ145336 |
| hCoV-19/Iraq/Samawa-49/2021 | Moderate | Live | - | MZ145292 |
| hCoV-19/Iraq/Samawa-50/2021 | Mild | Live | - | MZ145293 |
| hCoV-19/Iraq/Samawa- | Severe | Live | - | MZ145337 |

| | | | | |
|---|---|---|---|---|
| 51/2021 | | | | |
| hCoV-19/Iraq/Samawa-52/2021 | Mild | Live | - | MZ145295 |
| hCoV-19/Iraq/Samawa-53/2021 | Severe | Deceased | - | MZ366438 |
| hCoV-19/Iraq/Samawa-54/2021 | Mild | Live | - | MZ366439 |
| hCoV-19/Iraq/Samawa-55/2021 | Moderate | Live | - | MZ366440 |
| hCoV-19/Iraq/Samawa-56/2021 | Mild | Live | - | MZ366441 |
| hCoV-19/Iraq/Samawa-57/2021 | Mild | Live | - | MZ366442 |
| hCoV-19/Iraq/Samawa-58/2021 | Severe | Deceased | - | MZ366443 |
| hCoV-19/Iraq/Samawa-59/2021 | Severe | Deceased | - | MZ366444 |
| hCoV-19/Iraq/Samawa-60/2021 | Mild | Live | - | MZ366445 |
| hCoV-19/Iraq/Samawa-61/2021 | Moderate | Live | - | MZ366446 |
| hCoV-19/Iraq/Samawa-62/2021 | Moderate | Live | - | MZ366447 |
| hCoV-19/Iraq/Samawa-63/2021 | Mild | Live | - | MZ366448 |
| hCoV-19/Iraq/Samawa-64/2021 | Mild | Live | - | MZ366449 |
| hCoV-19/Iraq/Samawa-65/2021 | Moderate | Live | - | MZ366450 |
| hCoV-19/Iraq/Samawa-66/2021 | Mild | Live | - | MZ366451 |
| hCoV-19/Iraq/Samawa-67/2021 | Mild | Live | - | |
| hCoV-19/Iraq/Samawa-69/2021 | Mild | Live | - | MZ366452 |
| hCoV-19/Iraq/Samawa-71/2021 | Mild | Live | - | MZ366453 |
| hCoV-19/Iraq/Samawa-72/2021 | Moderate | Live | - | |
| hCoV-19/Iraq/Samawa-73/2021 | Moderate | Live | - | MZ366454 |
| hCoV-19/Iraq/Samawa-74/2021 | Moderate | Live | - | MZ366455 |
| hCoV-19/Iraq/Samawa-75/2021 | Moderate | Live | - | MZ366456 |
| hCoV-19/Iraq/Samawa-76/2021 | Moderate | Live | - | MZ366457 |
| hCoV-19/Iraq/Samawa-77/2021 | Moderate | Live | - | MZ366458 |
| hCoV-19/Iraq/Samawa-78/2021 | Moderate | Live | 27577* | |
